# Circulating microRNAs as potential biomarkers of hypertrophic cardiomyopathy phenotypic expression

**DOI:** 10.1101/2025.05.21.25325286

**Authors:** David Cordero Pereda, Patricia Fernandez San Jose, Rafael Moreno-Gómez Toledano, Sandra Perez-Rial, Luis Miguel Rincon Diaz, Pedro SeoaneZonjic, Sergio Fernandez Peñalver, Matias Morin, Natalia Sannikova, Simon Novo Flores, Patricia Rodriguez Solana, Juan Ranea, Miguel A Moreno-Pelayo, Jose Luis Zamorano Gomez

**Affiliations:** University Hospital Ramón y Cajal, Madrid, Spain; Instituto Ramón y Cajal de Investigación Sanitaria (IRYCIS), Madrid, Spain; CIBERCV - Spanish Network for Cardiovascular Research, Instituto de Salud Carlos III (ISCIII), Madrid, Spain; CIBERER - Center for Biomedical Network Research on Rare Diseases, Instituto de Salud Carlos III (ISCIII), Madrid, Spain; Universidad de Alcalá, Department of Surgery, Medical and Social Sciences, Area of Human Anatomy and Embryology, Alcalá de Henares 28871, Spain; Complejo Asistencial Universitario de Salamanca, IBSAL, Salamanca, Spain; Instituto de Investigación Biomédica de Salamanca (IBSAL), Universidad de Salamanca, Salamanca, Spain; University of Malaga, Malaga, Spain; Institute of Biomedical Research in Malaga and platform of nanomedicine, IBIMA Plataforma BIONAND, Malaga, Spain; Spanish National Bioinformatics Institute (INB/ELIXIR-ES), Instituto de Salud Carlos III (ISCIII), Madrid, Spain

**Keywords:** Hypertrophic cardiomyopathy (HCM), microRNAs (miRNAs), phenotypic expression, biomarkers, hypoxia-inducible factor-1 (HIF-1) pathway

## Abstract

**Background:** Hypertrophic cardiomyopathy (HCM) is characterized by significant variability in clinical presentation among patients with similar genetic backgrounds. The role of microRNAs (miRNAs) as epigenetic regulators in HCM remains underexplored. This pilot study aims to identify and characterize circulating miRNAs associated with different HCM phenotypes.

A cross-sectional study was conducted involving 28 participants: 10 with aggressive HCM, 10 with soft HCM, and 8 age- and sex-matched healthy controls. Circulating miRNAs were profiled using next-generation sequencing and quantitative real-time PCR. Differential expression analysis was performed to identify miRNAs associated with the aggressive phenotype.

**Results:** miR-16-5p, miR-17-5p, and let-7f-5p were identified as miRNAs significantly upregulated in the aggressive phenotype group compared to soft phenotypes and healthy controls. ROC analysis demonstrated statistically significant differences between aggressive HCM and softer forms with an area under the curve: miR-16-5p 0.833 (0.614 - 1), miR-17-5p 0.859 (0.648 – 1.00) and let-7f-5p 0.825 (0.62 – 1.00). After performing a target prediction analysis for the three miRNAs, the results were validated with three human transcriptomic datasets from control vs HCM patients. The analysis identified 34 quadruple coincidences across the four analyzed groups, and the enrichment analysis highlighted the hypoxia-inducible factor-1 (HIF-1) pathway.

**Conclusions:** miR-16-5p, miR-17-5p, and let-7f-5p appear to play a role in modulating the phenotypic expression of HCM. The potential use of these miRNAs as biomarkers for risk stratification and therapeutic targets to intervene in disease progression is promising. Validation in a wide clinical cohort is necessary to further elucidate the underlying mechanisms and therapeutic implications of miRNA dysregulation in HCM.

## BACKGROUND

Hypertrophic cardiomyopathy (HCM) is the most common inherited heart disease that primarily affects the cardiac muscle and constitutes a major cause of mortality and morbidity. The prevalence of HCM was classically established as 1:500, but the incorporation of genetic studies and refined imaging diagnosis has raised to 1:200. Numerous mutations have been described in more than 30 genes causing MCH, most of them in genes encoding sarcomeric proteins^1^. Consequently, there are potentially 750,000 HCM patients in the United States and 15-20 million worldwide ^2^.

HCM phenotypic expression and clinical repercussion are highly variable for the same genetic trait, and determinant factors have not been determined yet. Whereas some patients remain free of disease manifestations, and their life expectancy is not impaired, other patients have an extremely limited quality of life due to symptoms, HCM-related events, heart failure and sudden cardiac death. Current biomarkers reflect the consequences of severe established phenotypic expression and functional repercussion. While they provide prognostic information, they do not explain the cause of this variability in phenotypic expression. Among structural biomarkers left ventricular (LV) hypertrophy and geometry, left atrial volume and fibrosis are the most used for risk stratification ^3^. Functional biomarkers are represented by LV ejection fraction, myocardial mechanics and LV outflow obstruction ^4,5^. Natriuretic peptides and troponin detected in plasma or serum are used as unspecific markers on myocyte stress and injury, respectively ^6,7^. This phenotypic variability suggests the involvement of additional environmental factors, such as gender, hypertension, and physical activity ^8–12^, as well as epigenetic factors that influence disease progression. Among epigenetic regulators, DNA methylation, histone modification and non-coding RNAs, particularly microRNAs have been proposed as key modulators in multiple diseases, including cardiomyopathies ^13^.

MicroRNAs (miRNAs) are small, non-coding RNA molecules that are highly conserved across species and regulate gene expression by inhibiting translation or promoting messenger RNA (mRNA) degradation ^14^. miRNAs regulate post-transcriptional gene expression by binding to complementary sequences of target mRNA molecules. This interaction usually occurs in the 3‘untranslated region (3’ UTR) of the mRNA, which leads to the two previously mentioned consequences: degradation of the mRNA or inhibition of translation. While most miRNAs are intracellular, they can be found extracellularly in plasma and other fluids ^15–17^, a finding that has generated intense interest in extracellular miRNAs as biomarkers for a number of diseases, including myocardial infarction and heart failure ^18–21^. When miRNAs are overexpressed, they reduce the expression of their target genes through downregulation. In contrast, a reduced expression of miRNAs prevents the inhibition of target mRNAs, causing the corresponding genes to be overexpressed. As a result, miRNA expressivity strongly influences gene expression. By modulating gene expression and consequently the availability of specific proteins, miRNAs play a key role in the regulation of cellular processes such as growth, differentiation and apoptosis, and their dysregulation is linked to a variety of heart diseases. Certain differentially expressed miRNAs could be involved in phenotypic changes that progressively shift from a normal heart to cardiomyocyte hypertrophy, inflammation and fibrosis in HCM and may help explain why individuals with the same genetic mutations exhibit different clinical severities ^22–24^. Moreover, these biomarkers could contribute to discovering novel therapeutic targets, improving outcomes by intervening before the phenotypic expression of the disease.

Advances in genetic testing have identified a subgroup of HCM patients who are Genotype-positive–Phenotype-negative (G+P-). These individuals carry mutations associated with HCM but show no left ventricular hypertrophy or clinical symptoms ^25^. Identification of asymptomatic carriers has important implications for early risk stratification and personalized approaches to managing HCM, where factors like lifestyle changes and tailored surveillance can delay, or even prevent the onset of symptoms and cardiac adverse remodeling ^26,27^. In addition, certain miRNAs are known to regulate gene expression involved in cardiac hypertrophy and fibrosis, making them valuable tools for early detection ^28^. In summary, the identification of differentially expressed miRNAs that influence the severity of a patient’s phenotype could potentially enable clinicians to pre-identify patients at higher risk for a worse clinical course and adverse cardiac events. Moreover, these miRNAs could potentially become therapeutic targets to slow the progression to more aggressive HCM phenotypes.

In this study, we aim to compare the expression profiles of specific miRNAs across three well-defined groups with markedly different prognoses at the time of evaluation: healthy controls, patients with mild (soft) HCM, and those with severe HCM (aggressive) phenotypes.

## METHODS

The studies were conducted using peripheral blood samples from a subset of consecutive patients with HCM and variable expression of the disease and compared with paired controls. We selected 10 patients with HCM and severe signs/symptoms, 10 patients with HCM and minimum/no-expression of disease (both groups obtained from the Ramon y Cajal Biobank collection of Inherited Cardiac Diseases) and 8 sex and age matched controls (from the local Biobank collection of blood donors).

### Study population selection criteria

Patients with hypertrophic cardiomyopathy and **aggressive phenotype** with a known causative mutation in a cardiac sarcomere protein gene; massive hypertrophy (>30mm left-ventricular Wall thickness); late gadolinium enhancement in >20% of left-ventricular myocardium, assessed by cardiac MRI and severe (>50mmHg) left ventricular outflow tract obstruction or severe (>40ml/m^2^) left atrial dilation.
Patients with hypertrophic cardiomyopathy and **soft phenotype** with a known causative mutation in a cardiac sarcomere protein gene; familial history of clinically overt HCM with signs and symptoms; minimal or no hypertrophy (<15mm left-ventricular wall thickness); absence of late gadolinium enhancement and absence of significant (<15mmHg) left-ventricular outflow tract obstruction or left atrial dilation (28ml/m^2^).
Healthy **controls**: individuals matched by age and sex, without any personal or family history of cardiovascular diseases.

Genetic testing confirmed that all cases carried a pathogenic or likely pathogenic variant in a sarcomere gene, including MYH7, MYBPC3 and TNNT2. The pathogenic variants were classified according to ACMG criteria ^62^. Information concerning the genetic variants can be found in the **Table S1**.

### Sample collection and RNA extraction

Blood samples were collected in 4ml VACUETTE tubes (z serum sep clot activator), centrifuged at 2,500 rpm for 10 minutes. Serum was aliquoted and stored according to Biobank criteria (in anonymized tubes, with unique code and at -80C). Optimized RNA extraction protocols were used for human serum. For sequencing studies total RNA from all samples, including miRNA, was extracted from human serum using the “Total RNA Purification Kit” (Cat. #37500, Norgen Biotek Corp., Ontario, Canada) and for qRT-PCR using the “miRNeasy Serum Advanced Kit” (Cat. #217204, QIAGEN, Hilden, Germany) and always following the manufacturer’s instructions. Both long and small RNAs were kept in the same fraction and were referenced as total RNA. The quantity and quality of the total RNAs were evaluated using Agilent RNA 6000 Pico Chips (Cat. #5067-1513, Agilent Technologies, Santa Clara, CA, USA) by Agilent 2100 Bioanalyzer.

### Preparation of RNA libraries

Libraries generation and sequencing were performed at the Center for Cooperative Research in Biosciences (CIC bioGUNE; Bilbao) following the protocol included with the kit “NEXTflex™Small RNA-SeqKitv3,” (©Bioo Scientific Corp. Catalog #5132-05).. Briefly, 1 - 5 ng of total RNA was incubated for 2 minutes at 70°C, then 3’ 4N adenylated adapter and ligase enzyme were added and ligation was conducted by incubation of this mix overnight at 20°C. After excess 3’ adapter removal, 5%-adapter was added alongside with ligase enzyme and the mix was incubated at 20°C for 1 hour. The ligation product was used for the reverse transcription with the M-MuLV Reverse Transcriptase in a thermocycler for 30 min at 42°C and 10 min 90°C. Next, enrichment of the cDNA was performed using PCR cycling: RNA 2 min at 95°C; 25-42 cycles of 20 sec at 95°C, 30 sec at 60°C and 15 sec at 72°C; a final elongation of 2 min at 72°C and pause at 4°C. PCR products were resolved on 8% Novex TBE PAGE gels (Cat. #EC6215BOX, Thermo Fisher Scientific, Waltham, MA, USA), and fragments between 130 and 300 bp cut from the gel. Small RNAs were extracted from polyacrylamide gel using an adapted protocol, in which DNA from gel slices was dissolved in ddH2O overnight at room temperature and purified using NEXTFLEX Clean Up beads ^63^.

Amplified libraries concentration was determined with Qubit fluorometer using the Qubit®dsDNA HS assay kit (Cat. #Q32851, Thermo Fisher Scientific) and their size distribution was assessed running an aliquot on an Agilent Technologies 2100 Bioanalyzer, using an Agilent High Sensitivity DNA Chip (Cat. #5067-4626, Agilent Technologies).

### Sequencing and bioinformatic analysis

The sequencing was done using a High Throughput Sequencing, HiSeq 4000 machine, SR100 (1×100) (Illumina Technology). 20476188.46 reads were generated for all analyzed samples. The FASTQ files were pre-processed with BBTools software package (<u>BBMap download | SourceForge.net)</u> to remove adapters as described in the sequencing library documentation and to trim low-quality regions, with a quality lower than 26. Reads were aligned to the GRCh38 reference human genome assembly using Bowtie (v 1.0) ^62^ with restrictive parameters to avoid a possible multimapping effect: --seedmms 0 --maqerr 80 --seedlen 18 --all -m 7 --best --strata. Then, the aligned reads were analyzed with miRDeep ^29^ to identify the existing miRNA sequences in each sample. Once these miRNAs identifications were performed, a non-redundant miRNA reference was built by merging in a single FASTA file all the miRNAs identified in each sample and processing it with CD-HIT-EST (v4.8.1) ^30^ to remove redundant sequences.

These non-redundant predicted miRNA sequences were used as a reference with which to map the pre-processed reads with Bowtie2 (v2.2.9) ^31^ using the default options for single reads, and these alignments were used to obtain a count table to perform the miRNA differential expression analysis. This analysis was performed with the ExpHunter Suite Bioconductor package ^33–35^ using the algorithms DESeq2 and edgeR ^35^. In order to consider a miRNA as differentially expressed, we used as thresholds an adjusted p-value of false discovery rate (FDR) < 0.05 and an absolute value of log_2_ fold changes (log_2_ FC) > 1.

### Quantitative real-time polymerase chain reaction (qRT-PCR) assays

To validate the initial results of small RNA sequencing, qRT-PCR analysis was conducted on eight miRNAs (miR-16-5p, miR-17-5p, let-7f-5p, miR-30d-5p, miR-182-5p, miR-143-5p, miR-654-5p and miR-484). The RNA was reversely transcribed with the miRNA first strand cDNA synthesis (miRCURY LNA RT Kit, Cat. #339340, QIAGEN) according to the manufacturer’s instructions. The cDNA was quantitated using the SYBR miRNA RT-PCR kit (miRCURY LNA SYBR Green PCR Kit, Cat. #339345, QIAGEN) on a Roche LightCycler 480 II instrument (Roche Diagnostics, Basilea, Switzerland) under the following conditions: 2 minutes at 95°C, followed by 10 seconds at 95°C plus 60 seconds at 56°C for 45 cycles, then 5 seconds at 60°C and finally at 95°C. The relative expression levels of miRNAs were normalized to the mean of three invariantly expressed miRNAs like endogenous controls (miR-191-5p, miR-103a-3p and miR-93-5p) ^64^. Data analysis was performed using the second-derivative method. Relative fold change of miRNAs was calculated as 2^−ΔΔCt^. Results are expressed as median with interquartile range. For comparison purposes, the data for each group of patients were normalized to the median across all controls, as was done with sequencing data.

### Statistical analysis

Chi-square test (χ2) was used to compare quantitative variables in soft and aggressive phenotypes. Unpaired t-tests were used to compare continuous variables in soft and aggressive phenotypes when the normal distribution was confirmed by Shapiro-Wilk tests. Otherwise, Mann-Whitney U tests were used to compare two groups or one-way ANOVA, Kruskal-Wallis test with Dunn’s post-hoc analysis or two-way ANOVA with the Bonferroni post-test analysis was performed to compare three groups. The relative expression values of the aggressive phenotype miRNAs were normalized by their logarithmic correction. Subsequently, ROC curves were performed to identify individual predictive miRNAs of the aggressive phenotype. The statistical study was performed using the IBM SPSS Statistics for Windows software, version 26 (IBM Corp., Armonk, NY, USA) or the Prism version 8.0 (GraphPad Software, Inc., San Diego, CA, USA). All statistical test values below 0.05 were considered statistically significant.

### Target prediction and enrichment analysis

The bioinformatic analysis for the identification of miRNA target genes characteristic of the severity of the pathology (HC aggressive) was performed using FunRich open software (v.3.1.4). FunRich is a stand-alone software tool used for gene target prediction, functional enrichment, and interaction network analysis of genes and proteins ^65–67^. FunRich database includes Gene Ontology database, HPRD, Entrez Gene, and UniProt for the gene ontology annotations including biological process, cellular component, and molecular function. In addition, the mirTarbase and miRDB databases analyzed through the miRWalk web tool ^68^ were used. The core of miRWalk is the miRNA target site prediction with the random-forest-based approach software TarPmiR searching the complete transcript sequence including the 5’-UTR, CDS and 3’-UTR. All targets with a score greater than or equal to 0.90 were identified and all those targets with positive results were selected in at least two different databases.

Next, transcriptomic datasets related to HCM were identified on the National Library of Medicine official web (GEO DataSets, https://www.ncbi.nlm.nih.gov/gds). **GSE160997** (5 left ventricular free wall tissues from controls vs. 18 anterior septal tissues of HCM patients). **GSE180313** (7 cardiac tissues from control vs. 13 cardiac tissues of HCM patients), and **GSE36961** (39 cardiac tissues from control vs. 106 cardiac tissues of HCM patients). Data were analyzed with the GEO (Gene Expression Omnibus) 2R R language-based tool, and GEOquery ^69^, limma ^70^, and umap packages ^71^, identifying differentially expressed genes in patients with HCM that were overlapped with the targets previously identified in the predictive analysis of miRNAs (up-regulated miRNAs with down-regulated potential targets). Finally, all coincidental targets with all the four datasets (quadruple coincidence) were included in the enrichment analysis carried out. The Enrichr web tool was used to perform KEGG 2021 Human enrichment analysis ^72,73^. The ShinyGo 0.81 ^74^ web tool was used to perform GO biological process and KEGG enrichment analysis, and subsequently identify genes involved in the pathways of interest detected in the KEGG analysis.

## RESULTS

### Characteristics of the study population

We enrolled 28 participants, including 10 with an aggressive HCM-phenotype, 10 with a soft HCM-phenotype, and 8 age- and sex-matched healthy controls (see patients and methods). The myosin binding protein C (MYBPC3) gene mutation was the most prevalent (occurring in 80% and 50% in the aggressive and soft phenotypic group respectively), followed by myosin heavy chain (MYH7; 10% and 50% respectively) and troponin T (TNNC1, 10% only presented in the aggressive group. Patients were mostly female in the soft phenotype group, and men in the aggressive phenotype group. (80% vs. 10%. p. 0.002). Comparing both groups, patients with aggressive phenotype were more symptomatic (angina and dyspnea), had a high percentage of atrial fibrillation and ventricular arrhythmias, presence of late enhancement on MRI and increased ventricular thickness with a higher degree of mitral insufficiency. There were no significant differences in age, weight or previous history of hypertension between both groups of patients. All baseline characteristics of the patients are provided in **Table 1**.

**Table 1.** Patientś baseline characteristics. The data were expressed as a percentage for number of patients (no.) and as median and interquartile range (IQR) in quantitative variables. BMI, Body Mass Index; NYHA, New York Heart Association Functional Classification; ICD Implantable Cardioverter Defibrillator; MRI-LGE, Magnetic Resonance Imaging - Late gadolinium enhancement; LVOTG, Left Ventricular Outflow Tract Gradient; LVEF, Left Ventricular Ejection Fraction; MWT, maximal wall thickness; LV, Left Ventricular; LVEDD Left Ventricular End-Diastolic Diameter, LAV Left Atrial Volume.

| Variables | Aggressive phenotype<br>(n=10) | Soft phenotype<br>(n=10) | p-value<br>(0.05) |
| --- | --- | --- | --- |
| Age, years, median (IQR) | 43 [28.2 - 53.8] | 41.5 [35.2 - 46] | 0.733 |
| Women, no. (%) | 1 (10%) | 8 (80%) | **0.002** |
| BMI (Kg/m<sup>2</sup>), median (IQR) | 26.6 [26.2 - 29.8] | 21.6 [18.8 - 28] | 0.124 |
| Hypertension, no. (%) | 3 (30%) | 1 (10%) | 0.264 |
| Angina or dyspnea, no. (%) | 7 (70%) | 1 (10%) | **0.006** |
| Atrial fibrillation, no. (%) | 6 (60%) | 1 (10%) | **0.019** |
| Ventricular arrhythmia, no. (%) | 4 (40%) | 0 (0%) | **0.025** |
| ICD implant, no. (%) | 8 (80%) | 0 (0%) | **< 0.001** |
| MRI-LGE presence, no. (%) | 7 (70%) | 1 (10%) | **< 0.001** |
| LVOTG (mmHg), median (IQR) | 22.6 [8.8 - 33.2] | 8.1 [6.7 - 8.4] | 0.066 |
| LVEF, median (IQR) | 71.5 [59.3 - 74.2] | 70 [68.7 - 75.2] | 0.705 |
| Mitral E/E' ratio, median (IQR) | 12.4 [8.1 - 19.9] | 8.35 [7.1 - 10.28] | 0.102 |
| MWT (mm), median (IQR) | 31 [28 - 33.5] | 8.85 [8.03 - 9.98] | **< 0.001** |
| LV mass (g/m<sup>2</sup>), median (IQR) | 185 [151 - 264] | 71.65 [64.08 - 74.5] | **0.001** |
| LVEDD (mm), median (IQR) | 43 [35.75 - 48.75] | 40.5 [39.75 - 44.75] | 0.965 |
| LAV (ml/m<sup>2</sup>), median (IQR) | 49.68 [32.24 - 67.12] | 17.45 [13.31 - 21.59] | **0.041** |
| Burned-out phase, no. (%) | 1 (10%) | 0 (0%) | 0.305 |
| Medical therapy, no. (%) | 10 (100%) | 1 (10%) | **< 0.001** |
| Sepal myectomy, no. (%) | 2 (20%) | 0 (0%) | 0.136 |

### Identification of differentially expressed miRNAs in the aggressive phenotype

As shown in **Figure 1**, three different comparative analyses of data obtained by NGS sequencing were performed, identifying the differentially expressed miRNAs for each subgroup (aggressive vs. control, soft vs. control and aggressive vs. soft, **Figures 1A**, **1B** and **1C**, respectively). Subsequently, the overlapping analysis revealed eight miRNAs linked to the aggressive phenotype of HCM (**Figure 1D)**. A table with all the miRNAs detected is included in the **S*upplemental Material,* Table S2**.

**Figure 1.**
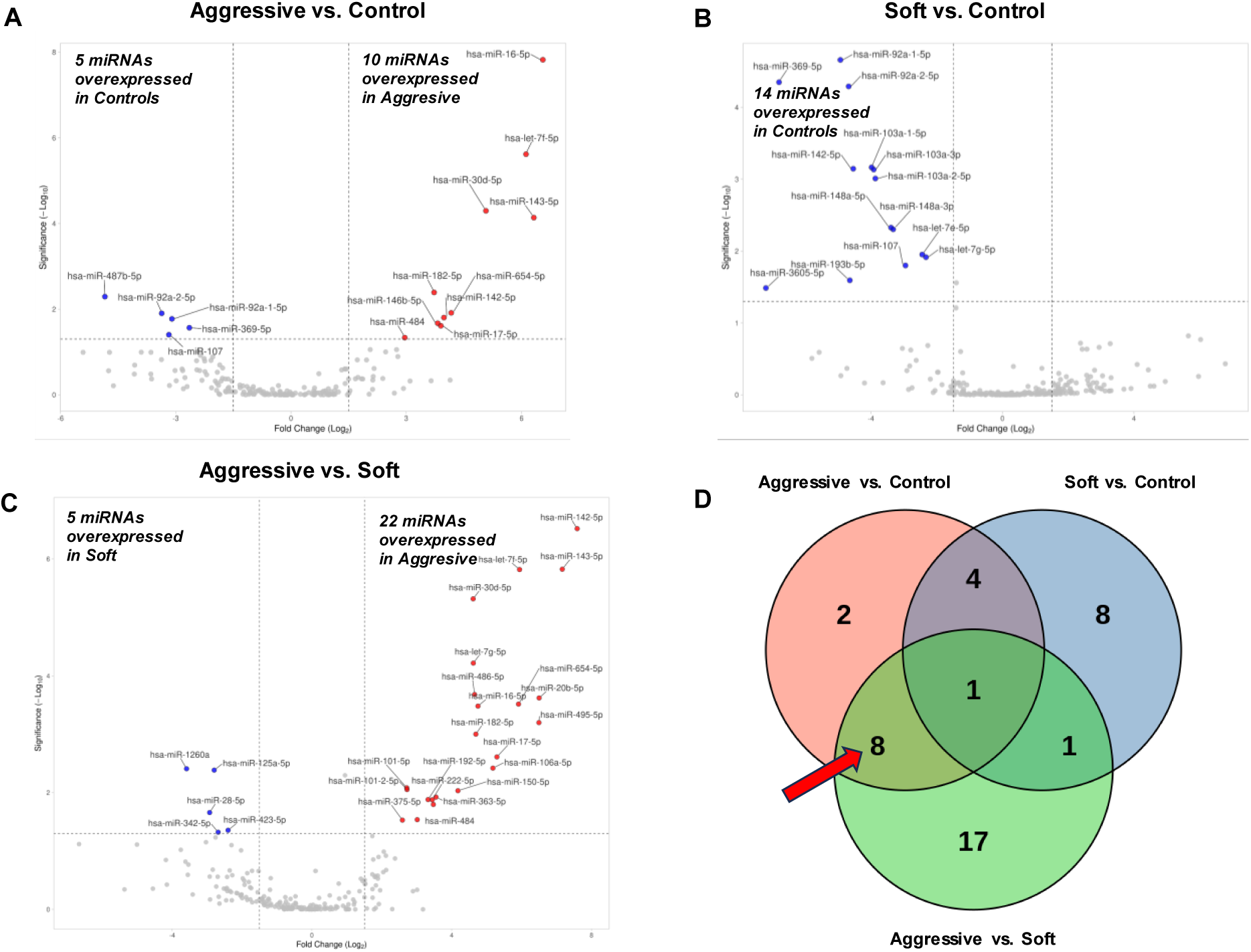
Identification of miRNAs characteristic of the aggressive phenotype. A-C) Representative volcano plots of the differentially expressed miRNA in the comparative analyses Aggressive vs. Control (A) Soft vs. Control (B), and Aggressive vs. Soft (C). Volcano plots highlighting statistically significant (-log_10_ (FDR) > 1.3) and differentially expressed (-1 < log_2_ FC > 1) miRNAs; D) Visualization of the representative differentially expressed miRNAs in the different studied subpopulations. The eight miRNAs overexpressed in the aggressive phenotype are marked with a red arrow.

As can be seen in **table 2**, the eight selected miRNAs are overexpressed in the aggressive phenotype.

**Table 2.** miRNAs linked to the aggressive phenotype of HCM. Note that FDR values (in bold) are only significant when being compared with the aggressive group.

|  | Aggressive vs. Control |  | Soft vs. Control |  | Aggressive vs Soft |  |
| --- | --- | --- | --- | --- | --- | --- |
| miRNA id | Mean FDR | Mean log<sub>2</sub>FC | Mean FDR | Mean log<sub>2</sub>FC | Mean FDR | Mean log<sub>2</sub>FC |
| hsa-miR-484 | **4.64E-02** | 2.960113 | 9.97E-01 | 0.186037 | **2.91E-02** | 3.012226 |
| hsa-miR-30d-5p | **5.06E-05** | 5.075065 | 6.17E-02 | -1.414430 | **4.83E-06** | 4.613683 |
| hsa-miR-16-5p | **1.51E-08** | 6.553497 | 9.98E-01 | -0.084529 | **3.30E-04** | 4.751758 |
| hsa-miR-182-5p | **4.07E-03** | 3.725194 | 9.89E-01 | 0.541443 | **9.98E-04** | 4.690753 |
| hsa-miR-17-5p | **2.45E-02** | 3.900114 | 7.29E-01 | 1.971222 | **2.45E-03** | 5.293947 |
| hsa-miR-654-5p | **1.22E-02** | 4.170136 | 9.62E-01 | 1.369718 | **3.05E-04** | 5.913171 |
| hsa-let-7f-5p | **2.42E-06** | 6.114646 | 9.87E-01 | 0.197872 | **1.52E-06** | 5.943200 |
| hsa-miR-143-5p | **7.31E-05** | 6.317892 | 9.99E-01 | -0.060692 | **1.50E-06** | 7.166220 |

Instructed by the miRNA-seq results, we further studied these eight miRNAs by qRT‒ PCR (***Supplemental Material***, **Figure S1**). The expression in the aggressive phenotype compared to the soft phenotype were statistically significant increased in **hsa-miR-16-5p** (1.884 [1.114 – 2.172] vs. 1.341 [0.9510 – 1.487], **\*** p = 0.0206), **hsa-miR-17-5p** (1.227 [0.9830 – 1.289] vs. 1.024 [0.8800 – 1.146], **\*** p = 0.0311) and **hsa-let-7f-5p** (2.013 [1.753 – 2.361] vs. 1.780 [1.430 – 2.033], **\*** p = 0.0311). Data are expressed as median with interquartile range (**Figure 2**).

**Figure 2.**
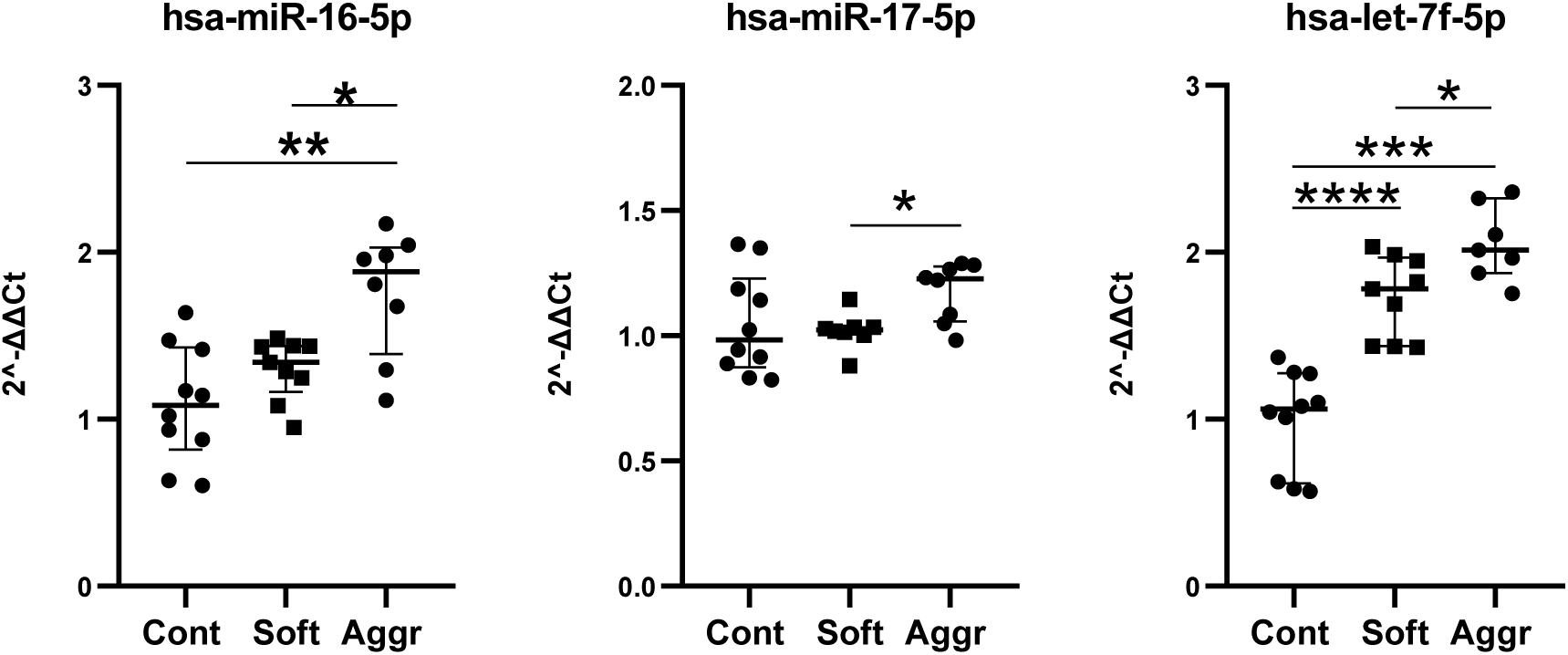
Expression levels of miRNAs with significant diagnostic value, quantified by qRT-PCR. n = 10 biological samples (per group). All experiments were performed at least in duplicate. Results were analyzed using the non-parametric Kruskal–Wallis test. Data are expressed as median with interquartile range. Cont, Control phenotype, Soft phenotype and Aggr, aggressive phenotype. * p < 0.05, ** p < 0.01, *** p < 0.001, **** p < 0.0001.

### Predictive models: ROC curves

The three previously selected miRNAs were individually analyzed using ROC curves, to identify the best individual biomarker that would allow distinguishing between the Aggressive and Soft phenotype (dichotomous variable: 0 Soft and 1 Aggressive). As can be seen in **Figure 3**, hsa-miR-16-5p, hsa-miR-17-5p and hsa-let-7f-5p show statistically significant expression, with an area under curve (AUC) (95% CI) of 0.833 (0.614-1.00), 0.859 (0.648-1.00), and 0.825 (0.62-1.00), respectively.

**Figure 3.**
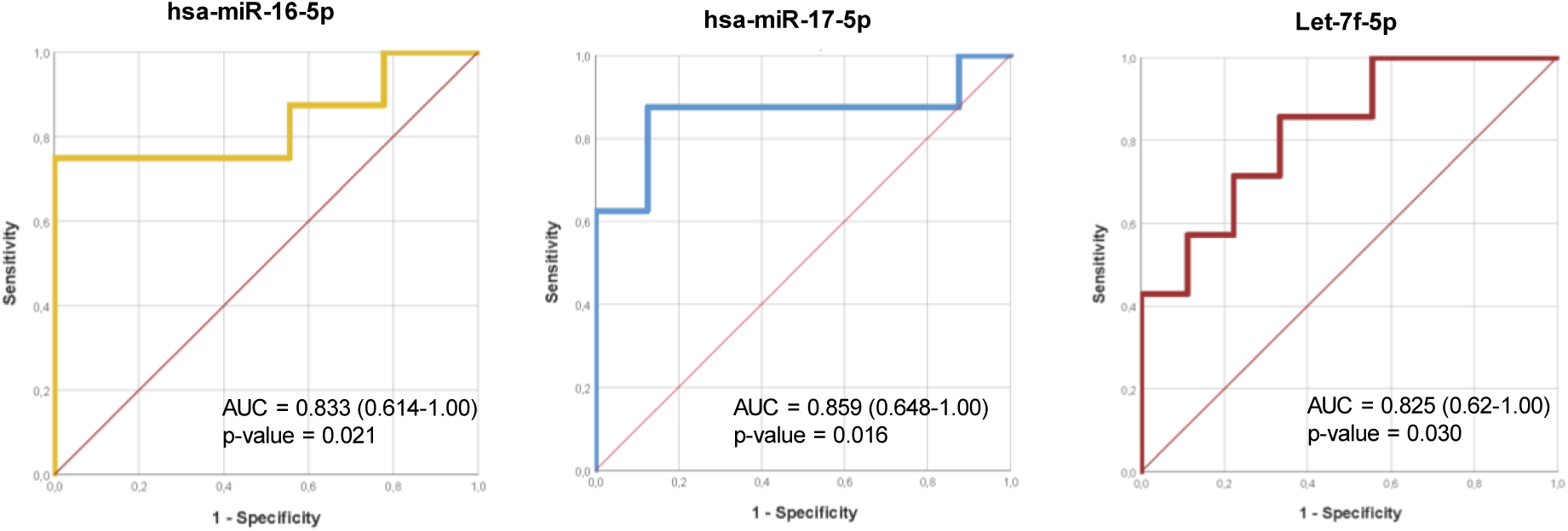
Visual representation of the discriminatory value study of miRNAs to identify patients with aggressive phenotype. Visual representation of the ROC curves of the three statistically significant miRNAs: hsa-miR-16-5p, hsa-miR-17-5p and hsa-let-7f-5p. AUC, area under curve. Cont, control. Aggr, aggressive. n = 10 biological samples (per group).

### Target prediction and enrichment analysis

After the identification of three miRNAs characteristic of the aggressive phenotypic expression of HCM, the predictive analysis of targets was performed. As can be seen in **Figure 4A**, 1,372 targets met the selection criteria (match in at least two predictive databases). All downregulated miRNAs in the HCM vs. control datasets were identified (**Figure 4B**) and the targets identified through the miRNA target prediction process were compared. As shown in **Figure 4C**, the results of the Venn diagram showed the quadruple matching of 34 genes. This result suggests that the phenotypic expression of aggressive HCM may be influenced by the regulation of these 34 genes, mediated by the expression levels of miR-16-5p, miR-17-5p, and let-7f-5p.

**Figure 4.**
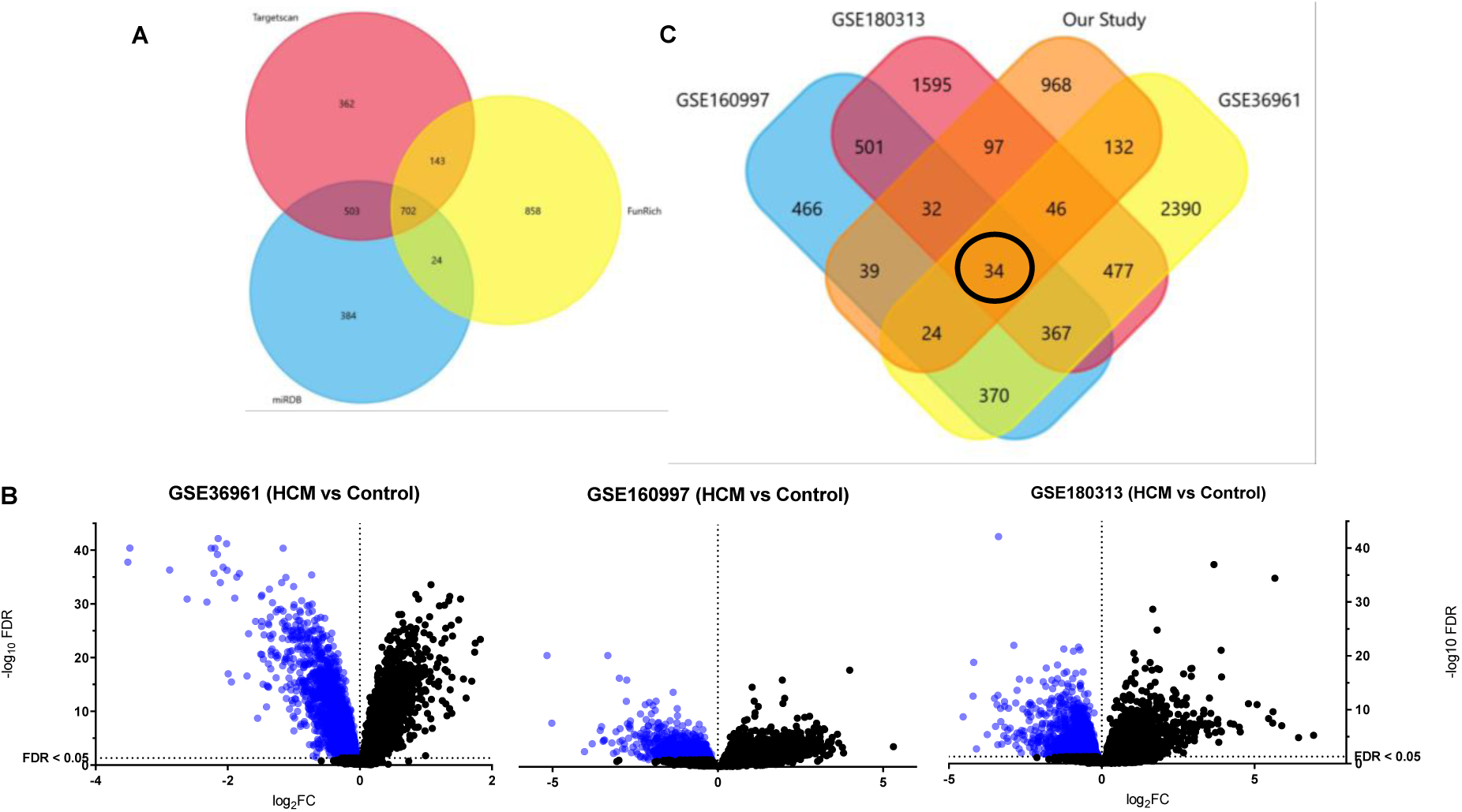
Target predictive analysis through GEO datasets. A) Target predictive analysis through Venn Diagram; B) Identification of HCM downregulated targets in the three selected datasets of HCM patients vs. Controls. The three selected datasets were GSE160997 (5 left ventricular free wall tissues from controls vs. 18 anterior septal tissues from HCM patients), GSE180313 (7 cardiac tissues from controls vs. 13 cardiac tissues from HCM patients), and GSE36961 (39 cardiac tissues from controls vs. 106 cardiac tissues from HCM patients. ; C) Venn diagram of the comparative study of the 3 human datasets vs. the dataset of predicted targets in this study. Note that in the center (black circle) the result corresponding to the quadruple coincidence has been encircled.

Subsequent enrichment analysis showed that, despite having a small number of genes, the GO Biological Process results determined several elements related to the cardiovascular system (**S*upplemental Material*, Figure S2A**), such as cardiac cell development, epithelial-mesenchymal transition, responses to growth factors, heart development and blood vessel morphogenesis. Successive KEGG 2021 Human enrichment analysis demonstrated the involvement of the hypoxia inducible factor-1 (HIF-1) signaling pathway, as well as genes involved in cardiac muscle contraction or even HCM itself (**Figure S2B**). By recreating the signaling pathways, we identified the genes that were within our 34 identified targets, observing the presence of STAT3 (Signal Transducer and Activator of Transcription 3), HIF-1α, TFRC, and EDN1 in the HIF-1 pathway (**Figure S2C**), SERCA2a and NHX in the cardiac muscle contraction pathway (**Figure S2D**), and finally SERCA2a and ET1 in the CMH pathway (**Figure S2E**).

## DISCUSSION

In this study, we have identified, for the first time, a differential expression profile of serum miRNAs associated with more aggressive HCM phenotype by analyzing their expression across three distinct groups with markedly different phenotypes and prognoses. The control group comprised anonymous donors from a biobank collection, selected based on the absence of personal or family history of cardiovascular disease, with no additional clinical data available. Consequently, baseline characteristic comparisons were performed exclusively between the soft and aggressive HCM phenotypic groups genetically confirmed. By analyzing the differential expression of miRNAs, we identified **miR-16-5p**, **miR-17-5p**, and **let-7f-5p** as key regulators potentially involved in the modulation of phenotypic expression in HCM. Their overexpression was correlated with an aggressive HCM phenotype, characterized by increased ventricular wall thickness, myocardial fibrosis, left atrial enlargement, left ventricular outflow tract gradient, and worse functional class.

Prior research has shown that miRNAs play a pivotal role in post-transcriptional regulation, significantly influencing the phenotype of HCM and magnitude of myocardial fibrosis. Derda et al. identified a molecular signature including miR-1, miR-21, miR-29a, miR-29b, miR-29c, miR-133a, miR-155, and miR-499, which may influence the severity of hypertrophy and fibrosis. Angelopoulos et al. highlighted miR-1, miR-29a, miR-133, miR-29a-3p, and miR-1-3p as potential biomarkers, with miR-29a playing a key role in distinguishing hypertrophic obstructive cardiomyopathy from non-obstructive HCM and other phenocopies ^22,23^. Fang et al. conducted an analysis of 84 circulating miRNAs in a cohort of eight patients with HCM, identifying 14 miRNAs with significantly elevated expression in individuals with diffuse myocardial fibrosis compared to control subjects. These 14 miRNAs were subsequently validated in an expanded group of 55 HCM patients. The study revealed reduced levels of miR-373-3p and miR-96-5p and elevated levels of 14 specific miRNAs (miR-18a-5p, miR-146a-5p, miR-30d-5p, miR-17-5p, miR-200a-3p, miR-19b-3p, miR-21-5p, miR-193-5p, miR-10b-5p, miR-15a-5p, miR-192-5p, miR-296-5p, miR-29a-3p, and miR-133a-3p) in HCM patients with diffuse myocardial fibrosis in contrast to those with non-diffuse fibrosis ^24^. Sucharov et al. investigated if circulating miRNAs could stratify disease stage in 57 patients with sarcomeric HCM, identifying differential expression of multiple miRNAs between carriers of sarcomeric mutations and healthy controls. This finding suggests that the presence of a sarcomeric variant, even with normal myocardial thickness, influences miRNA expression. However, the analyzed miRNAs did not distinguish between clinical and subclinical HCM ^29^. In contrast to our findings, most patients in the referenced study exhibited a non-aggressive hypertrophic cardiomyopathy phenotype with no major clinical differences among subjects that permitted an association between microRNA expression and phenotype severity.

We identified three miRNAs that were upregulated in the aggressive phenotype compared to both, the soft phenotype and healthy controls, which may help further discriminate the clinical relevance of differential miRNA expression.

The first one, miR-17-5p, constitutes a part of miR-17-92 cluster, which is known to regulate key cellular processes such as proliferation, growth, and apoptosis ^30^. These processes are essential in the development of cardiac hypertrophy and myocardial tissue remodeling, both of which are characteristic features of HCM ^24,31,32^. Fang et al. describes 14 circulating miRNAs, including miR-17-5p, which has been associated with diffuse myocardial fibrosis in hypertrophic cardiomyopathy. Notably, miR-17-5p exhibits a significant upregulation in patients with increased fibrosis burden, highlighting its potential as a non-invasive biomarker for disease progression and risk stratification ^24^. In relation to interactions with transcription factors and signaling pathways, it has been proposed that miR-17-5p interacts with transcription factors such as *STAT3* and *FASN*, which are implicated in hypertrophic responses and cardiovascular remodeling ^33^. The second one, miR-16-5p, promotes reticuloendothelial stress and oxidative stress in cardiac cells through regulating *ATF6*, suggesting that the inhibition of miR-16-5p has potential as a therapeutic approach to protect the heart against ER and oxidative stress-induced injury ^34,35^. In a rat ischemic model, knockdown of miR-16 demonstrated a cardioprotective role ^36^. The function of miR-16-5p in hypertrophic cardiomyopathy has not been previously described. Our findings are consistent with referenced studies in which the overexpression of miR-16-5p would lead to increased apoptosis, inflammation, and likely fibrosis, resulting in a more aggressive phenotype ^34–36^. Finally, let-7 miRNA was first discovered in *C. elegans* nematode and is highly conserved in human tissues ^37^. At least 12 distinct loci have been identified that encode different members of the let-7 family ^38^. Currently, let-7 family of miRNAs is considered one of the most extensively investigated. let-7 is expressed in all major types of cardiovascular cells. Previous studies have revealed the aberrant expression of let-7 members in cardiovascular diseases, such as heart hypertrophy *(let-7b/7c/7’s/miR-98*), cardiac fibrosis (*let-7c/7g*) and heart failure (*let-7b/7c/7d/7e/miR-98*) ^39,40^. A comprehensive review of the current scientific literature has not revealed any studies directly linking the miRNA let-7f-5p with hypertrophic cardiomyopathy. Furthermore, let-7f-5p has been observed to play a role in the regulation of T helper 17 (Th17) cell differentiation by inhibiting *STAT3*, suggesting its involvement in inflammatory processes ^39,40^.

Following the identification of genes related to the miRNAs of interest and their confirmation in the different human HCM cohorts analyzed, enrichment studies revealed pathways and genes of greater relevance in the pathophysiological context of HCM. KEGG analysis highlighted the HIF-1 signaling pathway as the most prominent, identifying several implicated genes such as *STAT3, HIF-1α, EDN1*, and *TFRC*. Evidence in the literature associates the HIF signaling pathway with cardiovascular remodeling, including cardiac hypertrophy and interstitial fibrosis ^41^. It is important to note that both the present study and the datasets analyzed to validate the miRNA targets involve patients with established HCM. This indicates that the relevance of the identified targets lies more in their association with disease progression rather than in predicting its onset.

The presence of HIF may be consistent with the understanding that cardiac muscle hypertrophy increases oxygen consumption and leads to tissue hypoxia ^41^. In the study by Chu et al., it was observed in a model of cultured neonatal rat cardiac myocytes that HIF-1α plays a key role in the development of cardiac hypertrophy in response to hypoxic stress ^42^. However, from the perspective of disease progression, the study by Shridhar et al. is particularly noteworthy ^43^. Using a murine HCM model with *MYBPC3* and *MYH6* mutations, the authors observed that the microvascular dysfunction occurring in HCM animals after myocardial hypertrophy is associated with an imbalance in the expression levels of HIF-1α and HIF-2α. Complementarily, and consistent with our findings, it has been reported that HIF-1α overexpression can attenuate cardiac dysfunction in the context of myocardial infarction ^44^. Therefore, in a scenario of downregulation mediated by the overexpression of inhibitory miRNAs, the dysregulation of HIF-1α could plausibly contribute to the negative progression of HCM ^45^_._

On the other hand, *STAT3* is a signaling molecule and transcription factor involved in various processes of cardiac protection. As illustrated in Figure S2C, STAT3 acts as a transcriptional regulator that upregulates HIF-1α mRNA and protein levels ^46^. Furthermore, it has been identified as a gene involved in developmental processes, critical for cardiac homeostasis, whose cooperation with HIF-1α regulates genes associated with developmental responses to oxidative stress ^47^. Overall, STAT3 mediates at multiple levels, including anti-oxidative, anti-apoptotic, and pro-angiogenic pathways. It has been shown to contribute to cardioprotection in various pathological contexts, such as cardiac hypertrophy, myocardial infarction, diabetic cardiomyopathy, and peripartum cardiomyopathy ^46^. As STAT3 is a direct target of miR-17-5p inhibiting its expression ^48^, the upregulation of miR-17-5p leads to the downregulation of STAT3, which in turn results in the reduced expression of HIF-1α, a key transcription factor involved in cellular responses to hypoxia and metabolic regulation.

Another highly interesting gene identified in Figure S2E is the cardiac isoform of the ATPase sarcoplasmic/endoplasmic reticulum Ca^2+^ transporting 2 (*ATP2A2* gene coding the SERCA2A protein), which was found to be involved in cellular signaling directly related to HCM. This finding aligns with the work of Wang et al. ^49^, who identified *ATP2A2* as the only gene differentially expressed when analyzing transcriptomic datasets from HCM patients. Reduced expression of this gene is associated with decreased calcium uptake by the sarcoplasmic reticulum in cardiomyocytes ^50^, suggesting that *ATP2A2* could be an important therapeutic target in various cardiovascular disorders due to the central role of the sarcoplasmic reticulum in myocardial contraction-relaxation coupling ^51^. This observation could also help explain the progression from HCM to heart failure, consistent with the findings of Yu et al. ^52^, who also identified *ATP2A2* as a key component in the interplay between hypoxia and HCM. The downregulation of *ATP2A2* (SERCA2a) in HCM leads to calcium dysregulation, which triggers hypertrophic remodeling, fibrosis, and metabolic dysfunction. The impaired relaxation increased fibrotic burden, and reduced ATP availability create a vicious cycle that promotes disease progression from HCM to heart failure. Given its central role in myocardial function, SERCA2a is an attractive therapeutic target for mitigating hypertrophy, fibrosis, and contractile dysfunction in HCM and other cardiovascular disorders ^53,54^.

Identification of miR-16-5p, miR-17-5p, and let-7f-5p in plasma as upregulated markers in aggressive HCM offers substantial implications for clinical management. The strong predictive values of these miRNAs, as demonstrated by ROC analysis, suggest that they could serve as valuable tools in clinical risk stratification as valuable indicators of disease progression, enabling more personalized monitoring and therapeutic strategies. However, in this study, we can only conclude that the identified upregulated miRNAs reflect a specific stage in the disease’s natural progression. Validation in a clinical cohort is required to determine their prognostic implications in the progression of hypertrophic cardiomyopathy severity. Our research group previously demonstrated in a prospective study the role of circulating miRNAs as prognostic biomarkers in patients presenting with MI ^21^ and functional tricuspid regurgitation ^55^.

From a therapeutic perspective, the inhibition or modulation of miR-16-5p, miR-17-5p, and let-7f-5p could represent a strategic approach to mitigating or delaying disease progression, particularly in high-risk individuals, as has already been proposed for disease as myocardial infarction and heart failure ^56–58^. The accumulating evidence has even encouraged experimental studies in animals and clinical trials focused on modulating miRNA expression using antisense therapy and anti-miRNA antibodies ^59,60^. As further studies validate these associations and elucidate the underlying mechanisms, these miRNAs could emerge as pivotal targets for the development of miRNA-based therapies for HCM, facilitating a more personalized approach to improve our understanding and management of this complex clinical condition ^61^.

Validation of these findings in an independent clinical cohort is necessary to confirm the prognostic value of the identified miRNAs. Future studies should incorporate patients at different stages of the disease to validate these miRNA signatures across varying genetic backgrounds, correlating the miRNA’s expression with clinical, biochemical and cardiac imaging variables. Additionally, mechanistic studies are essential to elucidate the precise molecular pathways through which these miRNAs affect disease progression, especially in relation to their interactions with other regulatory networks.

Thus, the present manuscript has demonstrated the potential impact that three miRNAs could have on the degree of phenotypic expression of CMH. Future studies will be necessary to deepen the paradigm, but the results have opened a new avenue in the diagnosis and prediction of MHC aggressiveness. In conclusion, miRNAs miR-16-5p, miR-17-5p, and let-7f-5p are significantly associated with the aggressive phenotypic expression of HCM. These miRNAs present promising potential as biomarkers for early diagnosis and risk stratification, as well as therapeutic targets to hinder disease progression. Further studies are essential to comprehensively elucidate their role in HCM and to advance the development of miRNA-based therapeutic interventions.

## DECLARATIONS

### Ethics approval and consent to participate

The study protocol was approved by the Clinical Research Ethics Committee of Hospital Ramón y Cajal (approval number 398) and was conducted in accordance with the principles outlined in the Declaration of Helsinki. Written informed consent was obtained from all participants or their legal guardians.

### Consent for publication

All participants provided written informed consent for the publication of the anonymized data derived from their biological samples and clinical records.

### Availability of data and materials

The sequencing data for this study have been deposited in the European Nucleotide Archive (ENA) at EMBL-EBI under accession number PRJEB86490 (https://www.ebi.ac.uk/ena/browser/view/PRJEB86490).

## Authors’ contributions

DCP, PFSJ, and RMG conceptualized the study, conducted experiments, and wrote the first draft. SPR, LMRD, PSZ, and SFP performed bioinformatic and statistical analyses. MM, NS, and SNF contributed to data interpretation and literature review. PRS, JR, and MAMP participated in experimental design and technical supervision. JLZG and MAMP supervised the project and critically revised the manuscript. All authors read and approved the final version.

## Acknowledgments

The authors would like to acknowledge Dr Laura García Bermejo and Dr. Leticia Olavarrieta Scappini for their valuable collaboration in the sequencing of miRNAs at the Centra Support Unit for translational Genomics, Instituto Ramón y Cajal de Investigación Sanitaria (IRYCIS). The author further express their gratitude to Sandra González Martín for her assistance with the statistical analysis and to Paz González Portilla for her support in sample collection.

## Authors’ information

- Dr. David Cordero Pereda is the coordinator of the Familial Cardiomyopathy Unit at Hospital Universitario Ramón y Cajal and a member of the CIBERCV network.
- Dr. Miguel A. Moreno-Pelayo is Head of the Department of Genetics at Hospital Universitario Ramón y Cajal and Principal Investigator at IRYCIS.
- Dr. José Luis Zamorano Gómez is Head of the Department of Cardiology at Hospital Universitario Ramón y Cajal and a leading expert in cardiovascular imaging and inherited cardiomyopathies.

The full list of author affiliations is provided on the title page of the manuscript.

## REFERENCES

1. Maron BJ, Maron MS, Maron BA, Loscalzo J. Moving Beyond the Sarcomere to Explain Heterogeneity in Hypertrophic Cardiomyopathy: JACC Review Topic of the Week. J Am Coll Cardiol. 2019;73:1978–1986.

2. Maron BJ, Rowin EJ, Maron MS. Hypertrophic Cardiomyopathy: New Concepts and Therapies. Annu Rev Med. 2022;73:363–375.

3. Hinojar R, Zamorano JL, Fernández-Méndez M, Esteban A, Plaza-Martin M, González-Gómez A, Carbonell A, Rincón LM, Nácher JJJ, Fernández-Golfín C. Prognostic value of left atrial function by cardiovascular magnetic resonance feature tracking in hypertrophic cardiomyopathy. Int J Cardiovasc Imaging. 2019;35:1055–1065.

4. Maron BJ, Desai MY, Nishimura RA, Spirito P, Rakowski H, Towbin JA, Rowin EJ, Maron MS, Sherrid MV. Diagnosis and Evaluation of Hypertrophic Cardiomyopathy: JACC State-of-the-Art Review. J Am Coll Cardiol. 2022;79:372–389.

5. Olivotto I, Cecchi F, Poggesi C, Yacoub MH. Patterns of disease progression in hypertrophic cardiomyopathy: an individualized approach to clinical staging. Circ Heart Fail. 2012;5:535–546.

6. Jansen M, Algül S, Bosman LP, Michels M, van der Velden J, de Boer RA, van Tintelen JP, Asselbergs FW, Baas AF. Blood-based biomarkers for the prediction of hypertrophic cardiomyopathy prognosis: a systematic review and meta-analysis. ESC Heart Fail. 2022;9:3418–3434.

7. Matthia EL, Setteducato ML, Elzeneini M, Vernace N, Salerno M, Kramer CM, Keeley EC. Circulating Biomarkers in Hypertrophic Cardiomyopathy. J Am Heart Assoc. 2022;11:e027618.

8. Maron BJ. Distinguishing hypertrophic cardiomyopathy from athlete’s heart physiological remodelling: clinical significance, diagnostic strategies and implications for preparticipation screening. Br J Sports Med. 2009;43:649–656.

9. Dimitrow PP, Czarnecka D, Strojny JA, Kawecka-Jaszcz K, Dubiel JS. Impact of gender on the left ventricular cavity size and contractility in patients with hypertrophic cardiomyopathy. Int J Cardiol. 2001;77:43–48.

10. Maron BJ, Casey SA, Hurrell DG, Aeppli DM. Relation of left ventricular thickness to age and gender in hypertrophic cardiomyopathy. Am J Cardiol. 2003;91:1195– 1198.

11. Page SP, Kounas S, Syrris P, Christiansen M, Frank-Hansen R, Andersen PS, Elliott PM, McKenna WJ. Cardiac myosin binding protein-C mutations in families with hypertrophic cardiomyopathy: disease expression in relation to age, gender, and long term outcome. Circ Cardiovasc Genet. 2012;5:156–166.

12. Pérez-Sánchez I, Romero-Puche AJ, García-Molina Sáez E, Sabater-Molina M, López-Ayala JM, Muñoz-Esparza C, López-Cuenca D, de la Morena G, Castro-García FJ, Gimeno-Blanes JR. Factors Influencing the Phenotypic Expression of Hypertrophic Cardiomyopathy in Genetic Carriers. Rev Esp Cardiol (Engl Ed). 2018;71:146–154.

13. Shi Y, Zhang H, Huang S, Yin L, Wang F, Luo P, Huang H. Epigenetic regulation in cardiovascular disease: mechanisms and advances in clinical trials. Signal Transduct Target Ther. 2022;7:200.

14. Correia de Sousa M, Gjorgjieva M, Dolicka D, Sobolewski C, Foti M. Deciphering miRNAs’ Action through miRNA Editing. Int J Mol Sci. 2019;20:6249.

15. Cortez MA, Bueso-Ramos C, Ferdin J, Lopez-Berestein G, Sood AK, Calin GA. MicroRNAs in body fluids--the mix of hormones and biomarkers. Nat Rev Clin Oncol. 2011;8:467–477.

16. Gilad S, Meiri E, Yogev Y, Benjamin S, Lebanony D, Yerushalmi N, Benjamin H, Kushnir M, Cholakh H, Melamed N, Bentwich Z, Hod M, Goren Y, Chajut A. Serum microRNAs are promising novel biomarkers. PLoS One. 2008;3:e3148.

17. Weber JA, Baxter DH, Zhang S, Huang DY, Huang KH, Lee MJ, Galas DJ, Wang K. The microRNA spectrum in 12 body fluids. Clin Chem. 2010;56:1733–1741.

18. Corsten MF, Dennert R, Jochems S, Kuznetsova T, Devaux Y, Hofstra L, Wagner DR, Staessen JA, Heymans S, Schroen B. Circulating MicroRNA-208b and MicroRNA-499 reflect myocardial damage in cardiovascular disease. Circ Cardiovasc Genet. 2010;3:499–506.

19. D’Alessandra Y, Devanna P, Limana F, Straino S, Di Carlo A, Brambilla PG, Rubino M, Carena MC, Spazzafumo L, De Simone M, Micheli B, Biglioli P, Achilli F, Martelli F, Maggiolini S, Marenzi G, Pompilio G, Capogrossi MC. Circulating microRNAs are new and sensitive biomarkers of myocardial infarction. Eur Heart J. 2010;31:2765–2773.

20. Fukushima Y, Nakanishi M, Nonogi H, Goto Y, Iwai N. Assessment of plasma miRNAs in congestive heart failure. Circ J. 2011;75:336–340.

21. Rincón LM, Rodríguez-Serrano M, Conde E, Lanza VF, Sanmartín M, González-Portilla P, Paz-García M, Del Rey JM, Menacho M, García Bermejo M-L, Zamorano JL. Serum microRNAs are key predictors of long-term heart failure and cardiovascular death after myocardial infarction. ESC Heart Fail. 2022;9:3367–3379.

22. Derda AA, Thum S, Lorenzen JM, Bavendiek U, Heineke J, Keyser B, Stuhrmann M, Givens RC, Kennel PJ, Schulze PC, Widder JD, Bauersachs J, Thum T. Blood-based microRNA signatures differentiate various forms of cardiac hypertrophy. Int J Cardiol. 2015;196:115–122.

23. Angelopoulos A, Oikonomou E, Vogiatzi G, Antonopoulos A, Tsalamandris S, Georgakopoulos C, Papanikolaou P, Lazaros G, Charalambous G, Siasos G, Vlachopoulos C, Tousoulis D. MicroRNAs as Biomarkers in Hypertrophic Cardiomyopathy: Current State of the Art. Curr Med Chem. 2021;28:7400–7412.

24. Fang L, Ellims AH, Moore X, White DA, Taylor AJ, Chin-Dusting J, Dart AM. Circulating microRNAs as biomarkers for diffuse myocardial fibrosis in patients with hypertrophic cardiomyopathy. J Transl Med. 2015;13:314.

25. Semsarian C, Ingles J, Maron MS, Maron BJ. New perspectives on the prevalence of hypertrophic cardiomyopathy. J Am Coll Cardiol. 2015;65:1249–1254.

26. Muresan ID, Agoston-Coldea L. Phenotypes of hypertrophic cardiomyopathy: genetics, clinics, and modular imaging. Heart Fail Rev. 2021;26:1023–1036.

27. Marian AJ, Braunwald E. Hypertrophic Cardiomyopathy: Genetics, Pathogenesis, Clinical Manifestations, Diagnosis, and Therapy. Circ Res. 2017;121:749–770.

28. Chiti E, Paolo MD, Turillazzi E, Rocchi A. MicroRNAs in Hypertrophic, Arrhythmogenic and Dilated Cardiomyopathy. Diagnostics (Basel). 2021;11:1720.

29. Sucharov CC, Neltner B, Pietra AE, Karimpour-Fard A, Patel J, Ho CY, Miyamoto SD. Circulating MicroRNAs Identify Early Phenotypic Changes in Sarcomeric Hypertrophic Cardiomyopathy. Circ Heart Fail. 2023;16:e010291.

30. Ambros V. The functions of animal microRNAs. Nature. 2004;431:350–355.

31. Bartel DP. MicroRNAs: target recognition and regulatory functions. Cell. 2009;136:215–233.

32. Liang LW, Hasegawa K, Maurer MS, Reilly MP, Fifer MA, Shimada YJ. Comprehensive Transcriptomics Profiling of MicroRNA Reveals Plasma Circulating Biomarkers of Hypertrophic Cardiomyopathy and Dysregulated Signaling Pathways. Circ Heart Fail. 2023;16:e010010.

33. Shi H, Li J, Song Q, Cheng L, Sun H, Fan W, Li J, Wang Z, Zhang G. Systematic identification and analysis of dysregulated miRNA and transcription factor feed-forward loops in hypertrophic cardiomyopathy. J Cell Mol Med. 2019;23:306–316.

34. Calderon-Dominguez M, Mangas A, Belmonte T, Quezada-Feijoo M, Ramos M, Toro R. Ischemic dilated cardiomyopathy pathophysiology through microRNA-16-5p. Rev Esp Cardiol (Engl Ed). 2021;74:740–749.

35. Toro R, Pérez-Serra A, Mangas A, Campuzano O, Sarquella-Brugada G, Quezada-Feijoo M, Ramos M, Alcalá M, Carrera E, García-Padilla C, Franco D, Bonet F. miR-16-5p Suppression Protects Human Cardiomyocytes against Endoplasmic Reticulum and Oxidative Stress-Induced Injury. Int J Mol Sci. 2022;23:1036.

36. Liu J, Sun F, Wang Y, Yang W, Xiao H, Zhang Y, Lu R, Zhu H, Zhuang Y, Pan Z, Wang Z, Du Z, Lu Y. Suppression of microRNA-16 protects against acute myocardial infarction by reversing beta2-adrenergic receptor down-regulation in rats. Oncotarget. 2017;8:20122–20132.

37. Lee RC, Feinbaum RL, Ambros V. The C. elegans heterochronic gene lin-4 encodes small RNAs with antisense complementarity to lin-14. Cell. 1993;75:843– 854.

38. Roush S, Slack FJ. The let-7 family of microRNAs. Trends Cell Biol. 2008;18:505– 516.

39. Su J-L, Chen P-S, Johansson G, Kuo M-L. Function and regulation of let-7 family microRNAs. Microrna. 2012;1:34–39.

40. Bao M-H, Feng X, Zhang Y-W, Lou X-Y, Cheng Y, Zhou H-H. Let-7 in cardiovascular diseases, heart development and cardiovascular differentiation from stem cells. Int J Mol Sci. 2013;14:23086–23102.

41. Sato T, Takeda N. The roles of HIF-1α signaling in cardiovascular diseases. J Cardiol. 2023;81:202–208.

42. Chu W, Wan L, Zhao D, Qu X, Cai F, Huo R, Wang N, Zhu J, Zhang C, Zheng F, Cai R, Dong D, Lu Y, Yang B. Mild hypoxia-induced cardiomyocyte hypertrophy via up-regulation of HIF-1α-mediated TRPC signalling. J Cell Mol Med. 2012;16:2022–2034.

43. Shridhar P, Glennon MS, Pal S, Waldron CJ, Chetkof EJ, Basak P, Clavere NG, Banerjee D, Gingras S, Becker JR. MDM2 Regulation of HIF Signaling Causes Microvascular Dysfunction in Hypertrophic Cardiomyopathy. Circulation. 2023;148:1870–1886.

44. Datta Chaudhuri R, Banik A, Mandal B, Sarkar S. Cardiac-specific overexpression of HIF-1α during acute myocardial infarction ameliorates cardiomyocyte apoptosis via differential regulation of hypoxia-inducible pro-apoptotic and anti-oxidative genes. Biochem Biophys Res Commun. 2021;537:100–108.

45. Luo Z, Tian M, Yang G, Tan Q, Chen Y, Li G, Zhang Q, Li Y, Wan P, Wu J. Hypoxia signaling in human health and diseases: implications and prospects for therapeutics. Signal Transduct Target Ther. 2022;7:218.

46. Harhous Z, Booz GW, Ovize M, Bidaux G, Kurdi M. An Update on the Multifaceted Roles of STAT3 in the Heart. Front Cardiovasc Med. 2019;6:150.

47. Grillo M, Palmer C, Holmes N, Sang F, Larner AC, Bhosale R, Shaw PE. Stat3 oxidation-dependent regulation of gene expression impacts on developmental processes and involves cooperation with Hif-1α. PLoS One. 2020;15:e0244255.

48. Liao X-H, Xiang Y, Yu C-X, Li J-P, Li H, Nie Q, Hu P, Zhou J, Zhang T-C. STAT3 is required for MiR-17-5p-mediated sensitization to chemotherapy-induced apoptosis in breast cancer cells. Oncotarget. 2017;8:15763–15774.

49. Wang X-Q, Yuan F, Yu B-R. Whole-Exome Sequencing Reveals Mutational Signature of Hypertrophic Cardiomyopathy. Int J Gen Med. 2023;16:4617–4628.

50. Lipskaia L, Chemaly ER, Hadri L, Lompre A-M, Hajjar RJ. Sarcoplasmic reticulum Ca(2+) ATPase as a therapeutic target for heart failure. Expert Opin Biol Ther. 2010;10:29–41.

51. Zarain-Herzberg A. Regulation of the sarcoplasmic reticulum Ca2+-ATPase expression in the hypertrophic and failing heart. Can J Physiol Pharmacol. 2006;84:509–521.

52. Yu H, Gu L, Du L, Dong Z, Li Z, Yu M, Yin Y, Wang Y, Yu L, Ma H. Identification and analysis of key hypoxia- and immune-related genes in hypertrophic cardiomyopathy. Biol Res. 2023;56:45.

53. Coppini R, Ferrantini C, Mugelli A, Poggesi C, Cerbai E. Altered Ca2+ and Na+ Homeostasis in Human Hypertrophic Cardiomyopathy: Implications for Arrhythmogenesis. Front Physiol. 2018;9:1391.

54. Angrisano T, Schiattarella GG, Keller S, Pironti G, Florio E, Magliulo F, Bottino R, Pero R, Lembo F, Avvedimento EV, Esposito G, Trimarco B, Chiariotti L, Perrino C. Epigenetic switch at atp2a2 and myh7 gene promoters in pressure overload-induced heart failure. PLoS One. 2014;9:e106024.

55. Hinojar R, Moreno-Gómez-Toledano R, Conde E, Gonzalez-Gomez A, García-Martin A, González-Portilla P, Fernández-Golfín C, García-Bermejo ML, Zaragoza C, Zamorano JL. Circulating miRNA in functional tricuspid regurgitation. Unveiling novel links to heart failure: A pilot study. ESC Heart Fail. 2024;11:2272–2286.

56. Condorelli G, Latronico MVG, Dorn GW. microRNAs in heart disease: putative novel therapeutic targets? Eur Heart J. 2010;31:649–658.

57. Song R, Dasgupta C, Mulder C, Zhang L. MicroRNA-210 Controls Mitochondrial Metabolism and Protects Heart Function in Myocardial Infarction. Circulation. 2022;145:1140–1153.

58. Täubel J, Hauke W, Rump S, Viereck J, Batkai S, Poetzsch J, Rode L, Weigt H, Genschel C, Lorch U, Theek C, Levin AA, Bauersachs J, Solomon SD, Thum T. Novel antisense therapy targeting microRNA-132 in patients with heart failure: results of a first-in-human Phase 1b randomized, double-blind, placebo-controlled study. Eur Heart J. 2021;42:178–188.

59. Hinkel R, Ramanujam D, Kaczmarek V, Howe A, Klett K, Beck C, Dueck A, Thum T, Laugwitz K-L, Maegdefessel L, Weber C, Kupatt C, Engelhardt S. AntimiR-21 Prevents Myocardial Dysfunction in a Pig Model of Ischemia/Reperfusion Injury. J Am Coll Cardiol. 2020;75:1788–1800.

60. Täubel J, Hauke W, Rump S, Viereck J, Batkai S, Poetzsch J, Rode L, Weigt H, Genschel C, Lorch U, Theek C, Levin AA, Bauersachs J, Solomon SD, Thum T. Novel antisense therapy targeting microRNA-132 in patients with heart failure: results of a first-in-human Phase 1b randomized, double-blind, placebo-controlled study. Eur Heart J. 2021;42:178–188.

61. Van Linthout S, Stellos K, Giacca M, Bertero E, Cannata A, Carrier L, Garcia-Pavia P, Ghigo A, González A, Haugaa KH, Imazio M, Lopes LR, Most P, Pollesello P, Schunkert H, Streckfuss-Bömeke K, Thum T, Tocchetti CG, Tschöpe C, van der Meer P, van Rooij E, Metra M, Rosano GMC, Heymans S. State of the art and perspectives of gene therapy in heart failure. A scientific statement of the Heart Failure Association of the ESC, the ESC Council on Cardiovascular Genomics and the ESC Working Group on Myocardial & Pericardial Diseases. Eur J Heart Fail. 2024;

62. Richards S, Aziz N, Bale S, Bick D, Das S, Gastier-Foster J, Grody WW, Hegde M, Lyon E, Spector E, Voelkerding K, Rehm HL, ACMG Laboratory Quality Assurance Committee. Standards and guidelines for the interpretation of sequence variants: a joint consensus recommendation of the American College of Medical Genetics and Genomics and the Association for Molecular Pathology. Genet Med. 2015;17:405–424.

63. Cordoba-Caballero J, Perkins JR, García-Criado F, Gallego D, Navarro-Sánchez A, Moreno-Estellés M, Garcés C, Bonet F, Romá-Mateo C, Toro R, Perez B, Sanz P, Kohl M, Rojano E, Seoane P, Ranea JAG. Exploring miRNA-target gene pair detection in disease with coRmiT. Brief Bioinform. 2024;25:bbae060.

64. Vasu MM, Koshy L, Ganapathi S, Jeemon P, Urulangodi M, Gopala S, Greeva P, Anitha A, Reethu S, Divya P, Shamla S, Sumitha K, Madhavan M, Vineeth CP, Kochumoni R, Harikrishnan S. Identification of novel endogenous control miRNAs in heart failure for normalization of qPCR data. Int J Biol Macromol. 2024;261:129714.

65. Fonseka P, Pathan M, Chitti SV, Kang T, Mathivanan S. FunRich enables enrichment analysis of OMICs datasets. J Mol Biol. 2021;433:166747.

66. Pathan M, Keerthikumar S, Ang C-S, Gangoda L, Quek CYJ, Williamson NA, Mouradov D, Sieber OM, Simpson RJ, Salim A, Bacic A, Hill AF, Stroud DA, Ryan MT, Agbinya JI, Mariadason JM, Burgess AW, Mathivanan S. FunRich: An open access standalone functional enrichment and interaction network analysis tool. Proteomics. 2015;15:2597–2601.

67. Mohashin P, Shivakumar K, David C, Riccardo A, Ching-Seng A, Philip A, Arsen O B, Alberto B-M, Giovanni C, Aled C, Federica C, Dolores DV, Jose Manuel F-P, Pedro F, Pamali F, Simona F, Yong Song G, An H, Esther N-H, Nunzio I, Kenneth K, Thomas K, Joana K, Igor V K, Tommaso L, Yaxuan L, Alicia L, Taral R L, Sayantan M, Francesca M, Anders Ø, Theocharis P, Tushar P, Héctor P, Stefano P, Simona P, Goran R, Felix R, Susmita S, Cristiana S, Allan S, Clotilde T, Martijn JC van H, Marca W, Joanne L W, Kening Z, Suresh M. A novel community driven software for functional enrichment analysis of extracellular vesicles data. Journal of extracellular vesicles [Internet]. 2017 [cited 2024 May 9];6. Available from: https://pubmed.ncbi.nlm.nih.gov/28717418/

68. Sticht C, De La Torre C, Parveen A, Gretz N. miRWalk: An online resource for prediction of microRNA binding sites. PLoS One. 2018;13:e0206239.

69. Davis S, Meltzer PS. GEOquery: a bridge between the Gene Expression Omnibus (GEO) and BioConductor. Bioinformatics. 2007;23:1846–1847.

70. Ritchie ME, Phipson B, Wu D, Hu Y, Law CW, Shi W, Smyth GK. limma powers differential expression analyses for RNA-sequencing and microarray studies. Nucleic Acids Res. 2015;43:e47.

71. McInnes L, Healy J, Melville J. UMAP: Uniform Manifold Approximation and Projection for Dimension Reduction [Internet]. 2018 [cited 2025 Feb 5];Available from: https://arxiv.org/abs/1802.03426

72. Kuleshov MV, Jones MR, Rouillard AD, Fernandez NF, Duan Q, Wang Z, Koplev S, Jenkins SL, Jagodnik KM, Lachmann A, McDermott MG, Monteiro CD, Gundersen GW, Ma’ayan A. Enrichr: a comprehensive gene set enrichment analysis web server 2016 update. Nucleic Acids Res. 2016;44:W90–97.

73. Chen EY, Tan CM, Kou Y, Duan Q, Wang Z, Meirelles GV, Clark NR, Ma’ayan A. Enrichr: interactive and collaborative HTML5 gene list enrichment analysis tool. BMC Bioinformatics. 2013;14:128.

74. Ge SX, Jung D, Yao R. ShinyGO: a graphical gene-set enrichment tool for animals and plants. Bioinformatics. 2020;36:2628–2629.

75. Van Driest SL, Jaeger MA, Ommen SR, Will ML, Gersh BJ, Tajik AJ, Ackerman MJ. Comprehensive analysis of the beta-myosin heavy chain gene in 389 unrelated patients with hypertrophic cardiomyopathy. J Am Coll Cardiol. 2004;44:602–610.

76. Cecconi M, Parodi MI, Formisano F, Spirito P, Autore C, Musumeci MB, Favale S, Forleo C, Rapezzi C, Biagini E, Davì S, Canepa E, Pennese L, Castagnetta M, Degiorgio D, Coviello DA. Targeted next-generation sequencing helps to decipher the genetic and phenotypic heterogeneity of hypertrophic cardiomyopathy. Int J Mol Med. 2016;38:1111–1124.

77. Williams N, Marion R, McDonald TV, Wang D, Zhou B, Eng LS, Um SY, Lin Y, Ruiter K, Rojas L, Sampson BA, Tang Y. Phenotypic variations in carriers of predicted protein-truncating genetic variants in MYBPC3: an autopsy-based case series. Cardiovasc Pathol. 2018;37:30–33.

78. Landstrom AP, Parvatiyar MS, Pinto JR, Marquardt ML, Bos JM, Tester DJ, Ommen SR, Potter JD, Ackerman MJ. Molecular and functional characterization of novel hypertrophic cardiomyopathy susceptibility mutations in TNNC1-encoded troponin C. J Mol Cell Cardiol. 2008;45:281–288.

79. Moolman JC, Brink PA, Corfield VA. Identification of a novel Ala797Thr mutation in exon 21 of the beta-myosin heavy chain gene in hypertrophic cardiomyopathy. Hum Mutat. 1995;6:197–198.

80. Niimura H, Bachinski LL, Sangwatanaroj S, Watkins H, Chudley AE, McKenna W, Kristinsson A, Roberts R, Sole M, Maron BJ, Seidman JG, Seidman CE. Mutations in the gene for cardiac myosin-binding protein C and late-onset familial hypertrophic cardiomyopathy. N Engl J Med. 1998;338:1248–1257.

81. García-Castro M, Reguero JR, Alvarez V, Batalla A, Soto MI, Albaladejo V, Coto E. Hypertrophic cardiomyopathy linked to homozygosity for a new mutation in the myosin-binding protein C gene (A627V) suggests a dosage effect. Int J Cardiol. 2005;102:501–507.

