## Supplementary material for "Circulating microRNAs as potential biomarkers of hypertrophic cardiomyopathy phenotypic expression": Suppl Material

**Table S1. Disease-causing variants in our cohort.** SNP ID: Single Nucleotide Polymorphism Identifier; ACMG American College of Medical Genetics and Genomics, P: pathogenic; LP: likely pathogenic; Het: heterozygous state.

| Subject | Phenotype | Gene | Exon | Nucleotide Change | Protein Change | Transcript | ACMG classification | Het. | References |
|---|---|---|---|---|---|---|---|---|---|
| 1 | Aggressive | *MYH7* | 30 | c.4066G>A | p.(Glu1356Lys) | NM_000257 | P (PP5, PP3, PM1, PM2) | Het | Van Driest, 2004[75] |
| 2 | Aggressive | *MYBPC3* | 16 | c.1624G>C | p.(Glu542Gln) | NM_000256 | P (PVS1, PP5, PM2) | Het | Ceconi, 2016[76] |
| 3 | Aggressive | *MYBPC3* | IVS 24 | c.2414-3C>G | p.? | NM_000256 | LP (PP3, PM2, PP1) | Het | *This study* |
| 4 | Aggressive | *MYBPC3* | 9 | c.884delT | p.(Phe295Serfs*5) | NM_000256 | P (PVS1, PP5, PM2) | Het | Williams, 2018[77] |
| 5 | Aggressive | *MYBPC3* | 17 | c.1624G>C | p.(Glu542Gln) | NM_000256 | P (PVS1, PP5, PM2) | Het | Ceconi, 2016[76] |
| 6 | Aggressive | *TNNC1* | 1 | c.23C>T | p.(Ala8Val) | NM_003280 | P (PS3, PP3, PP2) | Het | Landstrom, 2008[78] |
| 7 | Aggressive | *MYBPC3* | 19 | c.1808T>C | p.(Ile603Thr) | NM_000256 | LP (PP3, PM1, PM2) | Het | *This study* |
| 8 | Aggressive | *MYBPC3* | 26 | c.2724_2725delCTinsGCTGTA | p.(Tyr908*) | NM_000256 | LP (PVS1, PM2) | Het | *This study* |
| 9 | Aggressive | *MYBPC3* | 13 | c.1192A>T | p.(Lys398*) | NM_000256 | LP (PVS1, PM2) | Het | *This study* |
| 10 | Aggressive | *MYBPC3* | IVS 24 | c.2414-3C>G | p.? | NM_000256 | LP (PP3, PM2, PP1) | Het | *This study* |
| 11 | Soft | *MYH7* | 21 | c.2389G>A | p.(Ala797Thr) | NM_000257 | P (PM3, PP1, PM1, PP2, PM2, PM5, PP3) | Het | Moolman, 1995[79] |
| 12 | Soft | *MYH7* | 30 | c.4066G>A | p.(Glu1356Lys) | NM_000257 | P (PP5, PP3, PM1, PM2) | Het | Van Driest, 2004[75] |
| 13 | Soft | *MYBPC3* | 17 | c.1505G>A | p.(Glu375Lys) | NM_000256 | P (PS4, PM2, PM5, PM1, PP3) | Het | Niimura, 1998[80] |
| 14 | Soft | *MYBPC3* | 12 | c.1025T>A | p.(Val342Asp) | NM_170707 | LP (PM2, PM1, PP5, PP1) | Het | Garcia-Castro, 2005[81] |
| 15 | Soft | *MYBPC3* | 12 | c.1025T>A | p.(Val342Asp) | NM_000256 | LP (PM2, PM1, PP5, PP1) | Het | Garcia-Castro, 2005[81] |
| 16 | Soft | *MYH7* | 21 | c.2389G>A | p.(Ala797Thr) | NM_000257 | P (PM3, PP1, PM1, PP2, PM2, PM5, PP3) | Het | Moolman, 1995[79] |
| 17 | Soft | *MYH7* | 21 | c.2389G>A | p.(Ala797Thr) | NM_000257 | P (PM3, PP1, PM1, PP2, PM2, PM5, PP3) | Het | Moolman, 1995[79] |
| 18 | Soft | *MYBPC3* | 12 | c.1025T>A | p.(Val342Asp) | NM_170707 | LP (PM2, PM1, PP5, PP1) | Het | Garcia-Castro, 2005[81] |
| 19 | Soft | *MYBPC3* | 26 | c.2724_ 2725delCTinsGCTGTA | p.(Tyr908*) | NM_000256 | LP (PVS1, PM2) | Het | *This study* |
| 20 | Soft | *MYH7* | 21 | c.2389G>A | p.(Ala797Thr) | NM_000257 | P (PM3, PP1, PM1, PP2, PM2, PM5, PP3) | Het | Moolman, 1995[79] |

**Table S2. Table with all the miRNAs studied**. The eight differentially expressed miRNAs characteristic of the aggressive phenotype of HCM are marked in **gray.**

| | Aggressive vs Control | | Soft vs Control | | Aggressive vs Soft | |
|---|---|---|---|---|---|---|
| **miRNA id** | **Mean FDR** | **Mean $log_2FC$** | **Mean FDR** | **Mean $log_2FC$** | **Mean FDR** | **Mean $log_2FC$** |
| hsa-miR-16-5p | 1.51E-08 | 6.553498 | 9.98E-01 | -0.084529 | 3.30E-04 | 4.751759 |
| hsa-let-7f-5p | 2.42E-06 | 6.114646 | 9.87E-01 | 0.197873 | 1.52E-06 | 5.943200 |
| hsa-miR-30d-5p | 5.06E-05 | 5.075065 | 6.17E-02 | -1.414431 | 4.83E-06 | 4.613683 |
| hsa-miR-143-5p | 7.31E-05 | 6.317892 | 9.99E-01 | -0.060693 | 1.50E-06 | 7.166221 |
| hsa-miR-182-5p | 4.07E-03 | 3.725194 | 9.89E-01 | 0.541444 | 9.98E-04 | 4.690753 |
| hsa-miR-487b-5p | 5.10E-03 | -4.844929 | 8.69E-01 | -1.540114 | 7.07E-02 | -3.033308 |
| hsa-miR-654-5p | 1.22E-02 | 4.170137 | 9.62E-01 | 1.369719 | 3.05E-04 | 5.913172 |
| hsa-miR-92a-2-5p | 1.25E-02 | -3.365699 | 5.12E-05 | -4.683854 | 2.97E-01 | 1.884855 |
| hsa-miR-142-5p | 1.58E-02 | 3.980891 | 7.15E-04 | -4.543389 | 3.03E-07 | 7.589189 |
| hsa-miR-92a-1-5p | 1.69E-02 | -3.099234 | 2.19E-05 | -4.935126 | 2.30E-01 | 2.040009 |
| hsa-miR-146b-5p | 2.14E-02 | 3.818856 | 6.92E-01 | 1.549667 | 6.20E-01 | 1.502925 |
| hsa-miR-17-5p | 2.45E-02 | 3.900115 | 7.29E-01 | 1.971223 | 2.45E-03 | 5.293947 |
| hsa-miR-369-5p | 2.73E-02 | -2.645185 | 4.48E-05 | -6.801773 | 1.34E-01 | 2.375863 |
| hsa-miR-107 | 3.97E-02 | -3.178506 | 1.59E-02 | -2.950259 | 2.51E-01 | 1.726525 |
| hsa-miR-484 | 4.64E-02 | 2.960114 | 9.97E-01 | 0.186037 | 2.91E-02 | 3.012227 |
| hsa-let-7i-5p | 8.75E-02 | 2.061224 | 6.78E-01 | -1.329207 | 3.90E-01 | 1.828963 |
| hsa-miR-223-3p | 8.77E-02 | 2.747622 | 9.88E-01 | 0.156585 | 1.20E-01 | 2.115370 |
| hsa-miR-128-1-5p | 9.72E-02 | -2.417774 | 4.84E-01 | -1.960525 | 9.88E-01 | -0.290396 |
| hsa-miR-23b-5p | 9.93E-02 | 2.463042 | 5.58E-01 | 1.944857 | 3.64E-01 | 1.525359 |
| hsa-miR-432-5p | 1.01E-01 | -3.898225 | 3.31E-01 | -2.622817 | 8.60E-01 | -1.173604 |
| hsa-miR-493-5p | 1.01E-01 | -3.646167 | 9.45E-01 | -0.135061 | 2.09E-01 | -2.839162 |
| hsa-miR-548au-5p | 1.01E-01 | -5.413143 | 8.34E-01 | -1.183850 | 4.42E-01 | -4.554224 |
| hsa-miR-548b-5p | 1.01E-01 | -4.724946 | | | | |
| hsa-miR-128-2-5p | 1.19E-01 | -2.357824 | 4.84E-01 | -1.977405 | 9.90E-01 | -0.254404 |
| hsa-miR-495-5p | 1.27E-01 | 2.781214 | 9.04E-01 | -1.187101 | 6.33E-04 | 6.497821 |
| hsa-miR-106b-5p | 1.30E-01 | -2.035549 | 9.44E-01 | -0.835968 | 7.50E-01 | -0.911125 |
| hsa-miR-29a-5p | 1.42E-01 | -2.259799 | 9.98E-01 | 0.067168 | 2.40E-01 | -1.851581 |
| hsa-miR-3605-5p | 1.50E-01 | -3.372413 | | | | |
| hsa-let-7g-5p | 1.52E-01 | 1.724553 | 1.22E-02 | -2.330526 | 6.09E-05 | 4.618643 |
| hsa-miR-122-5p | 1.53E-01 | 1.965205 | 9.09E-01 | 1.062422 | 1.60E-02 | 3.474082 |
| hsa-miR-103a-2-5p | 1.57E-01 | -2.082801 | 9.78E-04 | -3.868958 | 3.62E-01 | 0.764284 |
| hsa-miR-323b-5p | 1.59E-01 | -2.457265 | 6.36E-01 | -1.493593 | 9.93E-01 | -0.323025 |
| hsa-miR-10b-5p | 1.62E-01 | -2.245892 | 9.82E-01 | 0.539332 | 7.94E-02 | -2.303518 |
| hsa-miR-103a-1-5p | 1.74E-01 | -2.055919 | | | | |
| hsa-miR-148a-3p | 2.00E-01 | -1.854303 | 4.97E-03 | -3.325214 | 9.56E-01 | -0.089166 |

| | | | | | | |
|---|---|---|---|---|---|---|
| hsa-miR-136-5p | 2.02E-01 | -4.069893 | | | | |
| hsa-miR-148a-5p | 2.36E-01 | -1.777623 | 4.71E-03 | -3.391765 | 9.20E-01 | 0.537733 |
| hsa-miR-192-5p | 2.42E-01 | 2.161576 | 9.99E-01 | 0.101168 | 1.32E-02 | 3.322595 |
| hsa-miR-24-1-5p | 2.48E-01 | 1.440273 | 9.88E-01 | 0.514935 | 2.65E-01 | 1.882643 |
| hsa-miR-24-2-5p | 2.48E-01 | 1.401079 | 9.85E-01 | 0.273502 | 2.68E-01 | 1.813510 |
| hsa-miR-24-3p | 2.48E-01 | 1.404859 | 9.84E-01 | 0.295648 | 2.79E-01 | 1.800782 |
| hsa-miR-29c-5p | 2.49E-01 | -2.834606 | 2.26E-01 | -3.045990 | 9.74E-01 | 0.391972 |
| hsa-miR-629-5p | 2.60E-01 | -2.449707 | 6.84E-01 | -1.183712 | 9.64E-01 | -0.567115 |
| hsa-miR-21-5p | 2.62E-01 | 1.316954 | 9.97E-01 | 0.220993 | 5.55E-02 | 1.726365 |
| hsa-miR-10399-5p | 2.70E-01 | 3.619800 | | | | |
| hsa-miR-1260b | 2.70E-01 | -3.019402 | 7.27E-01 | -1.466770 | 2.78E-01 | -2.428563 |
| hsa-miR-379-5p | 2.70E-01 | -2.377417 | 9.21E-01 | -1.182858 | 7.38E-01 | -0.740429 |
| hsa-miR-1304-5p | 2.71E-01 | -2.818615 | 3.96E-01 | -2.821508 | 9.81E-01 | -0.046195 |
| hsa-miR-1180-5p | 2.74E-01 | -4.749060 | | | | |
| hsa-miR-92b-5p | 2.87E-01 | -2.239327 | 7.23E-01 | -1.841919 | 9.85E-01 | 0.129899 |
| hsa-miR-185-5p | 2.96E-01 | -1.917037 | 9.84E-01 | -0.042968 | 9.76E-02 | -2.028726 |
| hsa-let-7a-5p | 3.21E-01 | 0.723015 | | | | |
| hsa-miR-378a-5p | 3.27E-01 | -4.078480 | 9.45E-01 | -1.584798 | 4.83E-01 | -2.348745 |
| hsa-miR-494-5p | 3.27E-01 | -3.649392 | 9.99E-01 | 0.424797 | 8.02E-01 | 0.363960 |
| hsa-miR-323a-5p | 3.39E-01 | -2.194439 | 6.77E-01 | 2.758159 | 1.71E-01 | -3.536984 |
| hsa-miR-454-5p | 3.73E-01 | 2.138996 | 1.94E-01 | 3.277879 | 2.95E-01 | -1.980291 |
| hsa-miR-486-5p | 3.91E-01 | 1.740217 | 2.50E-01 | -1.274594 | 2.09E-04 | 4.655308 |
| hsa-miR-548ap-5p | 3.96E-01 | -2.760284 | 9.92E-01 | 0.511571 | 7.97E-01 | -2.647899 |
| hsa-miR-99b-5p | 4.10E-01 | -1.784729 | 9.97E-01 | 0.169568 | 5.98E-01 | -1.370918 |
| hsa-miR-139-5p | 4.15E-01 | 2.389183 | 9.55E-01 | 1.416865 | 3.71E-01 | 1.517731 |
| hsa-miR-501-5p | 4.21E-01 | 1.834842 | 8.44E-01 | 1.112456 | 4.70E-01 | 1.949789 |
| hsa-miR-146a-5p | 4.23E-01 | 1.674603 | 1.91E-01 | 2.376539 | 5.77E-01 | 0.506667 |
| hsa-miR-378a-3p | 4.25E-01 | -3.628557 | 9.54E-01 | -0.343671 | 5.23E-01 | -2.160273 |
| hsa-miR-7-5p | 4.26E-01 | -1.710274 | 9.39E-01 | -0.486507 | 6.49E-01 | -1.430163 |
| hsa-miR-363-5p | 4.26E-01 | 1.866660 | 9.08E-01 | 1.443221 | 1.19E-02 | 3.549345 |
| hsa-miR-584-5p | 4.27E-01 | -1.638268 | 9.89E-01 | 0.328221 | 5.09E-01 | -1.553835 |
| hsa-miR-127-5p | 4.29E-01 | 1.498609 | 9.69E-01 | 1.310237 | 6.54E-01 | 0.776845 |
| hsa-miR-3177-5p | 4.53E-01 | 4.142310 | | | 8.11E-01 | 1.914656 |
| hsa-miR-625-5p | 4.53E-01 | 2.128865 | 7.66E-01 | 2.666110 | 8.87E-01 | 0.123337 |
| hsa-miR-6511b-5p | 4.57E-01 | -2.316677 | 7.98E-01 | -2.749599 | 1.00E+00 | 0.968579 |
| hsa-miR-3065-5p | 4.69E-01 | -2.986850 | 8.24E-01 | 1.717652 | 9.54E-02 | -3.755170 |
| hsa-miR-433-5p | 4.69E-01 | 3.646314 | 1.51E-01 | 5.655797 | 4.85E-01 | -2.422507 |
| hsa-miR-145-5p | 4.76E-01 | -1.820228 | 9.15E-01 | -0.135970 | 9.55E-01 | -0.811457 |
| hsa-miR-181a-5p | 4.76E-01 | -1.971144 | 9.30E-01 | 0.283937 | 5.86E-02 | -2.759720 |
| hsa-miR-196b-5p | 4.76E-01 | 3.098580 | 9.91E-01 | -1.090548 | 4.62E-01 | 3.009196 |
| hsa-miR-374a-5p | 4.76E-01 | -1.536735 | 7.46E-01 | -1.289408 | 9.98E-01 | 0.020769 |
| hsa-miR-769-5p | 5.33E-01 | -1.928921 | 9.27E-01 | -0.359030 | 6.91E-01 | -1.408756 |
| hsa-miR-140-5p | 5.45E-01 | 1.408568 | 9.69E-01 | 0.701208 | 1.03E-01 | 2.054067 |

| | | | | | | |
|---|---|---|---|---|---|---|
| hsa-miR-200c-5p | 5.92E-01 | -1.330151 | 8.42E-01 | -2.855806 | 1.00E+00 | 1.776921 |
| hsa-miR-1306-5p | 5.99E-01 | -1.886630 | 9.70E-01 | -1.246473 | 9.89E-01 | -0.776582 |
| hsa-miR-134-5p | 6.07E-01 | -3.163577 | 9.37E-01 | 2.082449 | 1.52E-01 | -3.621390 |
| hsa-miR-27a-5p | 6.10E-01 | -0.328708 | 9.91E-01 | 0.199534 | 4.62E-01 | -1.199996 |
| hsa-miR-331-5p | 6.14E-01 | -4.622777 | 9.86E-01 | -0.645159 | 9.33E-01 | -0.941568 |
| hsa-miR-30c-5p | 6.18E-01 | -1.251114 | 9.69E-01 | 0.172349 | 8.36E-01 | -0.783619 |
| hsa-miR-423-5p | 6.39E-01 | -1.143762 | 9.97E-01 | 0.344069 | 4.41E-02 | -2.400568 |
| hsa-miR-345-5p | 6.41E-01 | -2.705133 | 8.04E-01 | -1.499943 | 1.00E+00 | 0.253759 |
| hsa-miR-199a-5p | 6.44E-01 | -0.980855 | 9.99E-01 | 0.308621 | 2.19E-01 | -1.714522 |
| hsa-miR-191-5p | 6.52E-01 | 1.170806 | 9.89E-01 | 0.305458 | 1.27E-01 | 1.904623 |
| hsa-miR-125b-5p | 6.70E-01 | -1.616361 | 9.70E-01 | -0.037743 | 8.05E-01 | -1.171785 |
| hsa-miR-25-5p | 6.73E-01 | 0.342517 | 8.04E-01 | 1.021820 | 9.21E-01 | -0.301332 |
| hsa-miR-148b-5p | 6.82E-01 | -1.321449 | 9.69E-01 | 0.896882 | 3.39E-01 | -1.807253 |
| hsa-miR-450b-5p | 6.87E-01 | -2.067417 | 5.78E-01 | 3.303243 | 7.77E-02 | -5.009731 |
| hsa-miR-26a-5p | 6.93E-01 | 0.610815 | 4.94E-01 | -1.211331 | 3.27E-01 | 1.448724 |
| hsa-miR-4284 | 7.11E-01 | -1.642163 | 9.17E-01 | -1.637342 | 1.00E+00 | 0.512621 |
| hsa-miR-431-5p | 7.11E-01 | 2.926492 | 4.54E-01 | 2.800171 | 6.17E-01 | -0.761632 |
| hsa-miR-335-5p | 7.21E-01 | 0.970809 | 2.31E-01 | 2.428277 | 9.27E-01 | -0.697553 |
| hsa-miR-424-5p | 7.21E-01 | -2.800453 | 9.84E-01 | 1.015738 | 2.75E-01 | -2.926156 |
| hsa-miR-19b-2-5p | 7.24E-01 | -1.093921 | 7.01E-01 | -1.486979 | 8.19E-01 | 1.042545 |
| hsa-miR-421 | 7.45E-01 | -0.486967 | 9.70E-01 | 1.162490 | 4.31E-01 | -2.434447 |
| hsa-miR-99a-5p | 7.45E-01 | 2.123332 | 5.58E-01 | 2.678248 | 7.82E-01 | -0.939318 |
| hsa-miR-30a-5p | 7.45E-01 | -0.994819 | 9.70E-01 | -0.612883 | 9.90E-01 | 0.165675 |
| hsa-miR-19b-1-5p | 7.61E-01 | -0.805499 | 6.84E-01 | -1.371004 | 7.51E-01 | 1.219089 |
| hsa-miR-19a-5p | 7.87E-01 | -1.935961 | 9.85E-01 | 0.158974 | 7.18E-01 | -1.285142 |
| hsa-miR-942-5p | 7.90E-01 | -0.851940 | 8.60E-01 | 1.879808 | 2.60E-01 | -2.280755 |
| hsa-miR-652-5p | 7.90E-01 | 1.136269 | 7.32E-01 | 1.918265 | 9.27E-01 | -0.666341 |
| hsa-miR-574-5p | 7.98E-01 | 1.534162 | 9.62E-01 | 1.533044 | 9.63E-01 | 0.187473 |
| hsa-miR-222-5p | 8.02E-01 | 0.894238 | 9.98E-01 | 0.361978 | 1.33E-02 | 3.442767 |
| hsa-let-7e-5p | 8.08E-01 | -0.103199 | 1.12E-02 | -2.441333 | 8.87E-01 | 0.689952 |
| hsa-miR-199b-5p | 8.15E-01 | 0.589565 | 9.85E-01 | 0.097932 | 2.12E-01 | 2.122257 |
| hsa-miR-941 | 8.25E-01 | -1.705424 | 9.89E-01 | 0.667179 | 5.30E-01 | -2.136123 |
| hsa-miR-409-5p | 8.26E-01 | 0.843789 | 8.71E-01 | 1.265255 | 8.00E-01 | -0.455747 |
| hsa-miR-106a-5p | 8.31E-01 | -0.289572 | 2.38E-01 | -2.789844 | 3.81E-03 | 5.179777 |
| hsa-miR-370-5p | 8.33E-01 | -1.564359 | 9.84E-01 | 1.265649 | 4.06E-01 | -2.739155 |
| hsa-miR-320a-5p | 8.35E-01 | -0.664906 | | | | |
| hsa-miR-1307-5p | 8.35E-01 | -1.616849 | 9.70E-01 | 0.879211 | 6.57E-01 | -1.243499 |
| hsa-miR-1260a | 8.40E-01 | -0.907780 | 4.27E-01 | 1.198138 | 3.90E-03 | -3.589591 |
| hsa-miR-15a-5p | 8.42E-01 | 1.034884 | 7.31E-01 | 1.582186 | 9.15E-01 | -0.557076 |
| hsa-miR-98-5p | 8.48E-01 | -0.714589 | 9.45E-01 | -0.863630 | 6.38E-01 | 0.200006 |
| hsa-miR-4433b-5p | 8.49E-01 | 0.826403 | 7.10E-01 | 1.699255 | 6.61E-01 | -0.611203 |
| hsa-miR-29b-1-5p | 8.53E-01 | -1.449185 | 9.98E-01 | -0.516939 | 8.51E-01 | -0.036757 |
| hsa-miR-29b-2-5p | 8.53E-01 | -1.503262 | 9.70E-01 | -1.181297 | 1.00E+00 | 1.237195 |

| | | | | | | |
|---|---|---|---|---|---|---|
| hsa-miR-3173-5p | 8.53E-01 | -1.225636 | 9.83E-01 | 0.517809 | 8.98E-01 | -1.059454 |
| hsa-miR-340-5p | 8.53E-01 | 1.231120 | 9.69E-01 | 1.134141 | 8.70E-01 | 0.887016 |
| hsa-miR-320c | 8.57E-01 | -1.062664 | 9.70E-01 | -0.377688 | 9.98E-01 | -0.170788 |
| hsa-miR-4732-5p | 8.57E-01 | -1.687068 | 8.98E-01 | -1.214521 | 9.68E-01 | 0.675902 |
| hsa-miR-342-5p | 8.58E-01 | -0.780647 | 6.65E-01 | 2.027253 | 4.77E-02 | -2.684019 |
| hsa-miR-27b-5p | 8.61E-01 | -0.283109 | 9.54E-01 | -0.276514 | 1.00E+00 | -0.017963 |
| hsa-miR-96-5p | 8.62E-01 | -0.029801 | 9.70E-01 | -1.025528 | 6.30E-01 | 1.734498 |
| hsa-miR-1843 | 8.74E-01 | 1.776982 | | | | |
| hsa-miR-150-5p | 8.87E-01 | 0.611990 | 9.86E-01 | -0.286448 | 9.28E-03 | 4.178174 |
| hsa-miR-30e-5p | 8.88E-01 | 0.460052 | 9.69E-01 | -0.554570 | 2.99E-01 | 1.397840 |
| hsa-miR-485-5p | 8.99E-01 | 0.743423 | 9.99E-01 | 0.255154 | 8.25E-01 | 1.198890 |
| hsa-miR-101-5p | 9.00E-01 | 0.668218 | 9.27E-01 | 0.999687 | 8.28E-03 | 2.712038 |
| hsa-miR-500a-5p | 9.10E-01 | 1.508544 | 6.81E-01 | 2.246807 | 6.54E-01 | -1.426593 |
| hsa-miR-186-5p | 9.12E-01 | 0.112036 | 9.84E-01 | 0.566598 | 9.82E-01 | 0.360008 |
| hsa-miR-215-5p | 9.13E-01 | 0.924860 | 8.24E-01 | 2.066906 | 8.97E-01 | 0.218400 |
| hsa-miR-487a-5p | 9.13E-01 | 2.689862 | | | 5.05E-03 | 0.944311 |
| hsa-miR-374b-5p | 9.21E-01 | -1.054554 | 9.45E-01 | 1.641009 | 4.47E-01 | -2.461683 |
| hsa-miR-221-5p | 9.22E-01 | -0.430986 | 9.92E-01 | -0.317274 | 9.99E-01 | 0.095258 |
| hsa-miR-125a-5p | 9.23E-01 | -0.623656 | 2.31E-01 | 2.569766 | 4.12E-03 | -2.797765 |
| hsa-miR-20a-5p | 9.28E-01 | 0.605484 | 9.12E-01 | -1.394962 | 4.89E-01 | 1.359813 |
| hsa-miR-450a-5p | 9.35E-01 | -0.733486 | 9.95E-01 | -0.985729 | 9.99E-01 | 1.002296 |
| hsa-miR-93-5p | 9.40E-01 | 0.013365 | 9.16E-01 | 0.681314 | 9.98E-01 | -0.085720 |
| hsa-miR-330-5p | 9.50E-01 | -1.874610 | 9.56E-01 | 1.935634 | 2.56E-01 | -3.553247 |
| hsa-miR-22-5p | 9.52E-01 | -0.172933 | 9.98E-01 | 0.219578 | 6.12E-01 | 0.948925 |
| hsa-miR-15b-5p | 9.55E-01 | -0.518269 | 9.94E-01 | -0.308953 | 9.99E-01 | 0.071440 |
| hsa-miR-543 | 9.66E-01 | 1.144577 | 9.69E-01 | 1.304010 | 9.99E-01 | -0.065731 |
| hsa-miR-1-3p | 9.67E-01 | 2.033276 | 9.20E-01 | 2.778869 | 9.86E-01 | -0.575590 |
| hsa-miR-11400 | 9.67E-01 | 0.238077 | | | | |
| hsa-miR-224-5p | 9.67E-01 | 0.988101 | 9.97E-01 | 0.283073 | 9.17E-01 | 0.728024 |
| hsa-miR-328-5p | 9.67E-01 | -1.249900 | 9.69E-01 | 1.304028 | 5.05E-01 | -2.315149 |
| hsa-miR-375-5p | 9.67E-01 | -0.096085 | 9.69E-01 | 1.047575 | 2.96E-02 | 2.587669 |
| hsa-miR-6803-5p | 9.67E-01 | 0.373582 | 8.75E-01 | 1.605002 | 5.98E-01 | -0.470152 |
| hsa-miR-155-5p | 9.68E-01 | -0.980150 | 9.98E-01 | -0.185634 | 9.98E-01 | -0.294280 |
| hsa-miR-20b-5p | 9.68E-01 | 0.554785 | 6.79E-01 | -3.633819 | 2.39E-04 | 6.503120 |
| hsa-miR-576-5p | 9.68E-01 | -0.954848 | 9.70E-01 | 1.464003 | 7.34E-01 | -1.588845 |
| hsa-miR-144-5p | 9.68E-01 | -0.062657 | 9.86E-01 | -0.513806 | 8.36E-01 | 0.034234 |
| hsa-miR-126-5p | 9.69E-01 | 0.244343 | 6.81E-01 | 1.238068 | 9.27E-01 | -0.570302 |
| hsa-miR-3613-5p | 9.70E-01 | 1.026899 | 9.20E-01 | 1.623795 | 9.72E-01 | -0.549550 |
| hsa-miR-4446-5p | 9.70E-01 | -1.095811 | 9.84E-01 | 0.410766 | 8.17E-01 | -1.280470 |
| hsa-miR-26b-5p | 9.73E-01 | -0.276138 | 9.88E-01 | 0.113351 | 2.20E-01 | -1.526870 |
| hsa-miR-151b | 9.76E-01 | -0.168443 | 8.71E-01 | -0.687140 | 1.40E-01 | 1.752650 |
| hsa-miR-1301-5p | 9.77E-01 | 1.110131 | 6.80E-01 | 2.863958 | 7.77E-01 | -1.359633 |
| hsa-miR-197-5p | 9.78E-01 | -0.399048 | 8.18E-01 | 1.696730 | 4.33E-01 | -1.740059 |

| | | | | | | |
|---|---|---|---|---|---|---|
| hsa-miR-320e | 9.78E-01 | -0.652344 | 8.18E-01 | 1.830017 | 5.56E-01 | -2.013563 |
| hsa-miR-504-5p | 9.78E-01 | -1.853181 | | | | |
| hsa-miR-370-3p | 9.79E-01 | 1.104672 | 9.77E-01 | 1.640717 | 9.83E-01 | -0.635262 |
| hsa-miR-382-5p | 9.79E-01 | 0.999082 | 6.22E-01 | 2.590273 | 4.19E-01 | -1.768237 |
| hsa-miR-339-5p | 9.84E-01 | 0.944485 | 9.53E-01 | 1.727430 | 9.74E-01 | -0.616869 |
| hsa-miR-532-5p | 9.87E-01 | -0.583979 | 7.79E-01 | 1.692253 | 3.22E-01 | -1.938542 |
| hsa-miR-30b-5p | 9.88E-01 | -0.688035 | 9.69E-01 | 1.118062 | 6.83E-01 | -1.506380 |
| hsa-miR-194-5p | 9.92E-01 | 0.773924 | 7.08E-01 | 2.623173 | 7.50E-01 | -1.425710 |
| hsa-miR-320b | 9.92E-01 | -0.096477 | 9.98E-01 | 0.273056 | 1.00E+00 | 0.000678 |
| hsa-miR-660-5p | 9.96E-01 | 0.615328 | 8.91E-01 | 1.747919 | 7.57E-01 | -1.710249 |
| hsa-miR-28-5p | 9.96E-01 | -0.187386 | 2.18E-01 | 2.938669 | 2.20E-02 | -2.928752 |
| hsa-miR-101-2-5p | 9.97E-01 | 0.236489 | 9.21E-01 | 0.811184 | 8.87E-03 | 2.719787 |
| hsa-miR-183-5p | 9.97E-01 | -0.166357 | 6.38E-01 | 2.104232 | 3.79E-01 | -1.927744 |
| hsa-miR-23a-5p | 9.98E-01 | -0.087051 | 9.69E-01 | -0.678035 | 3.14E-01 | 1.509216 |
| hsa-let-7c-5p | 9.98E-01 | 0.024600 | 6.13E-01 | 1.766516 | 2.92E-01 | -1.179519 |
| hsa-miR-130b-5p | 9.99E-01 | -0.120412 | 9.70E-01 | 0.876390 | 8.81E-01 | -0.873011 |
| hsa-miR-181b-5p | 9.99E-01 | 0.347612 | 9.10E-01 | 1.713841 | 8.30E-01 | -0.678967 |
| hsa-miR-18a-5p | 9.99E-01 | 0.020151 | 9.80E-01 | 1.088270 | 9.83E-01 | -0.578403 |
| hsa-miR-193a-5p | 9.99E-01 | -0.190047 | 9.98E-01 | 0.508887 | 9.60E-01 | -0.617222 |
| hsa-miR-195-5p | 9.99E-01 | 0.844797 | 9.99E-01 | 0.094492 | 6.79E-01 | 1.178948 |
| hsa-miR-23c | 9.99E-01 | 1.468648 | 9.70E-01 | -0.708650 | 4.88E-01 | 2.890897 |
| hsa-miR-3615 | 9.99E-01 | -0.418816 | 9.70E-01 | 0.111048 | 9.26E-01 | -0.379995 |
| hsa-miR-483-5p | 9.99E-01 | -0.260667 | 9.70E-01 | 1.108159 | 8.47E-01 | -1.363445 |
| hsa-miR-505-5p | 9.99E-01 | -0.155824 | 5.34E-01 | 2.562910 | 2.16E-01 | -2.354455 |
| hsa-miR-885-5p | 9.99E-01 | -0.131879 | 9.99E-01 | -0.146898 | 1.00E+00 | -0.795998 |
| hsa-miR-1972 | 1.00E+00 | 0.591666 | 8.92E-01 | 1.202345 | 9.90E-01 | -0.308556 |
| hsa-miR-100-5p | 1.00E+00 | -1.177544 | 9.05E-01 | 2.101478 | 6.17E-01 | -1.883501 |
| hsa-miR-10a-5p | 1.00E+00 | 0.051436 | 9.01E-01 | 1.434052 | 6.85E-01 | -1.228089 |
| hsa-miR-1322 | 1.00E+00 | -0.309660 | 6.05E-01 | -2.198736 | 4.50E-01 | 1.731009 |
| hsa-miR-502-5p | 1.00E+00 | 0.977765 | 9.32E-01 | 1.842451 | 9.24E-01 | -0.449627 |

**Figure S1. Expression levels of miRNAs candidates by qRT-PCR**. Data are expressed as median with interquartile range. Cont, Control phenotype, Soft phenotype and Aggr, aggressive phenotype. * $p < 0.05$, ** $p < 0.01$, *** $p < 0.001$, **** $p < 0.0001$.

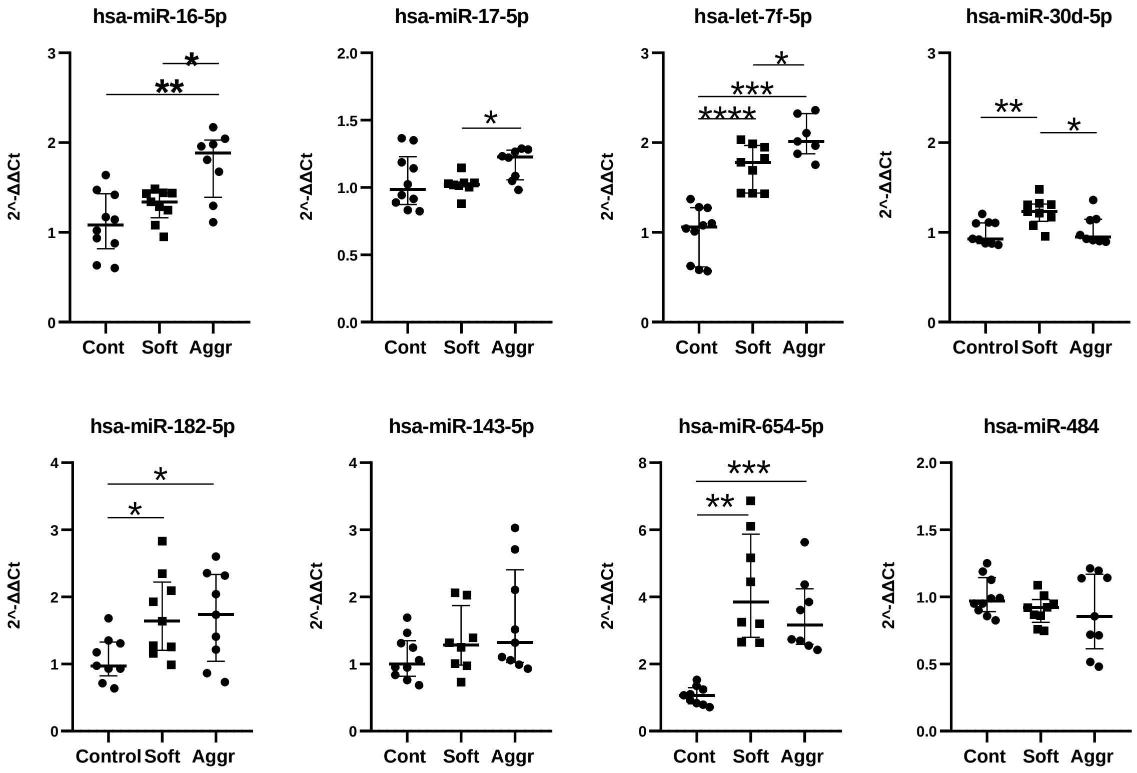

**Figure S2. Complementary analysis of validated targets.** A) Gene Ontology (GO) Biological Process enrichment analysis highlighting processes related to cardiovascular pathologies; B) KEGG 2021 Human pathway analysis showing that the most significant result corresponds to the HIF-1 signaling pathway; C-E) Visual representation of the most relevant signaling pathways and the implicated genes identified through the quadruple coincidence analysis: HIF-1 signaling pathway (C), cardiac muscle contraction (D), and hypertrophic cardiomyopathy (E).

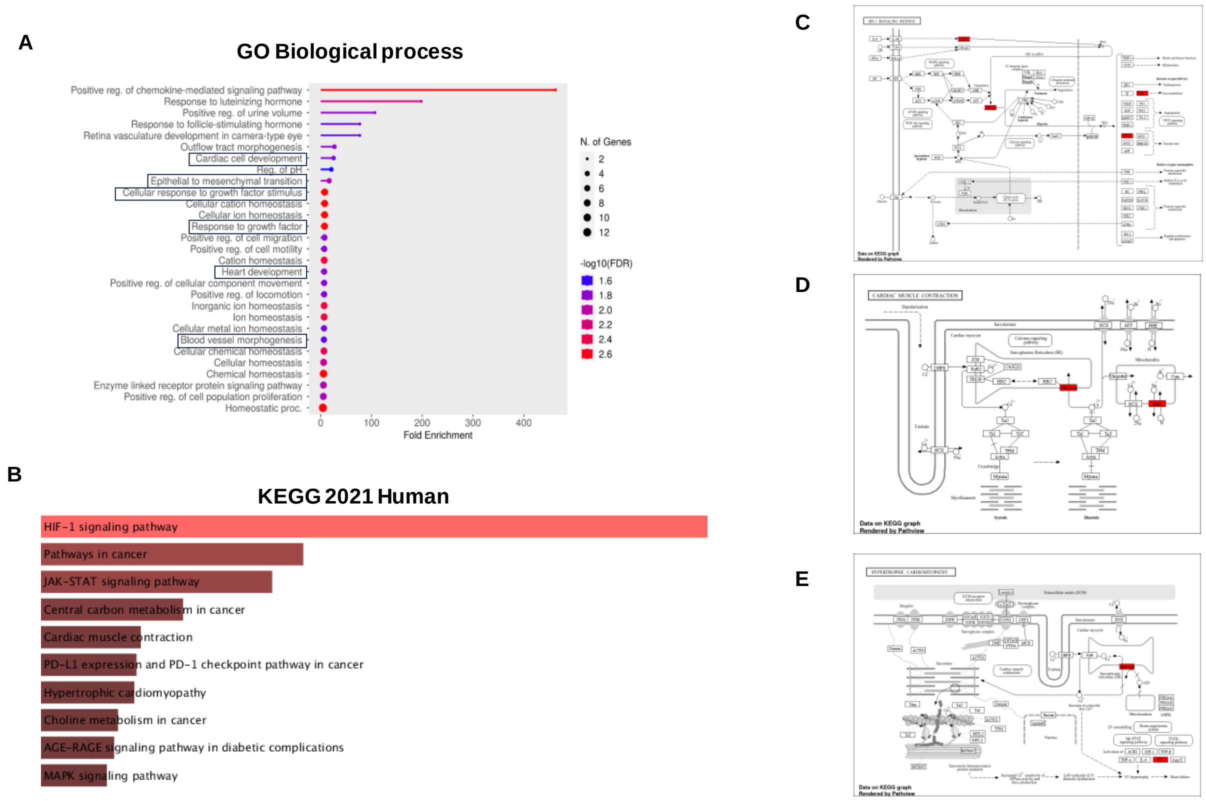
